# Effectiveness of Hand Hygiene Interventions in Healthcare Facilities in Low- and Middle-Income Countries (LMICs): A Systematic Review

**DOI:** 10.64898/2026.07.28.26359185

**Authors:** Fred Tusabe, Ryan Cronk, Fred Twinomugisha, Mackline Ninsiima, Darcy Anderson, Lucy K Tantum, Kenneth Kobba, Reuben Kiggundu, Dathan Byonanebye, Judith Nanyondo Semanda, Francis Kakooza

**Affiliations:** Infectious Diseases Institute, Makerere University, Kampala, Uganda; Department of Environmental Sciences and Engineering, Gillings School of Global Public Health, University of North Carolina at Chapel Hill, 135 Dauer Drive, CB#7431, Chapel Hill, North Carolina 27599, United States; School of Public Health, Makerere University

**Keywords:** Hand hygiene, Infection prevention and control, WASH in health-care facilities, barriers and enablers

## Abstract

**Background:** Hand hygiene is a core component of infection prevention and control, yet evidence on the effectiveness of hand hygiene interventions in health-care facilities in low-income and middle-income countries has not been comprehensively synthesized alongside implementation barriers and enablers. We assessed the effectiveness of hand hygiene interventions in these settings and examined implementation conditions shaping success.

**Methods:** We conducted a systematic review drawing on a University of North Carolina evidence map of environmental health services in low-income and middle-income country health-care facilities, which searched PubMed, Scopus, and Global Health and was supplemented by hand-searching. Hand-hygiene-specific records were restricted to peer-reviewed English-language studies published between Jan 1, 2015, and Nov 28, 2025; backward citation chasing identified additional eligible records. Random-effects meta-analysis was done for studies with extractable pre-intervention and post-intervention hand hygiene compliance data. Other outcomes were synthesized using effect-direction methods. Barriers and enablers were synthesized thematically. The protocol was registered with PROSPERO, CRD420251252831.

**Findings:** We screened 656 records and included 57 studies from 32 low-income and middle-income countries. Fifteen studies contributed to meta-analysis. Hand hygiene interventions were associated with higher post-intervention than pre-intervention compliance (pooled risk ratio 1·45, 95% CI 1·20–1·74), with substantial heterogeneity (I²=98%). Most studies used non-randomized designs. Multimodal WHO-style strategies and system-change interventions focused on alcohol-based hand rub availability, placement, or related infrastructure were the most common intervention categories. Effect-direction synthesis suggested favorable effects for compliance and system-related outcomes, whereas evidence for reductions in health-care-associated infections was less consistent. Common barriers included supply shortages, weak audit and data systems, staffing and workload pressures, and WASH infrastructure constraints; common enablers included training, monitoring and feedback, leadership support, reliable supplies, and implementation support.

**Conclusion:** Hand hygiene interventions can improve observed compliance in low-income and middle-income country health-care facilities, but sustained gains depend on system supports that make hand hygiene feasible at the point of care. Programs should pair training and behavior-change strategies with reliable supplies, functional WASH infrastructure, audit-and-feedback routines, leadership accountability, and protected implementation time.

## Introduction

Healthcare-associated infections (HAIs) remain a major, largely preventable cause of morbidity, mortality, and prolonged hospitalization, with a disproportionate burden in low- and middle-income countries (LMICs) [1]. A recent meta-analysis of 87 studies conducted in LMICs (2000-2024) reported an overall HAI prevalence of 22%, with substantially higher prevalence in the WHO South-East Asia Region (37%) and in low-income countries (37%) [2]. In contrast, earlier global syntheses have consistently shown lower endemic HAI prevalence in high-income settings (approximately 7% of hospitalized patients) compared with LMICs (about 15%) [1]. This disparity likely reflects differences in baseline risk, surveillance capacity, staffing, and infrastructure constraints, and the feasibility of implementing core infection prevention and control (IPC) measures in many LMIC settings[3].

Hand hygiene is a foundation of IPC because it interrupts transmission of pathogens via healthcare workers’ hands during routine care [4]. The World Health Organization (WHO) identifies hand hygiene as central to preventing infection and notes that improved hand hygiene can avert a substantial proportion of avoidable HAIs [4]. Nevertheless, hand hygiene compliance is often suboptimal, particularly in settings with constrained resources. WHO summaries indicate markedly lower hand hygiene compliance in low-income compared with high-income settings in intensive care contexts (e.g., 9% vs 65% across studies up to 2018) [5]. These gaps are clinically important because hand hygiene improvements have been associated with reductions in infection outcomes in real-world programs, although effect sizes and certainty vary by design and outcome ascertainment. For example, in a landmark hospital-wide program implemented in a high-income country setting, sustained hand hygiene compliance over time coincided with reductions in healthcare-associated infections and methicillin-resistant Staphylococcus aureus transmission across [6].

In LMICs, persistent structural constraints can limit both baseline hand hygiene opportunities and the durability of improvement strategies [7]. The WHO/UNICEF Joint Monitoring Program (JMP) report on water, sanitation, and hygiene (WASH) in health care facilities highlights that, globally, 3.4 billion people used facilities lacking basic hygiene services in 2023 [3]. Using the JMP service ladder, “basic hygiene” requires functional hand hygiene facilities at points of care and near toilets; in 2022, only 57% of facilities had basic hygiene services, while 9% had no hygiene service, and 516 million people used facilities with no hygiene service at the point of care [3]. Facility-level observational evidence is consistent with these system constraints; alcohol-based hand rub was available at the point of care for only 56% of opportunities, while soap and water were available for 10 % [8]. Together, these data underscore why hand hygiene improvement in LMIC health care facilities must be considered both a behavioral and a systems challenge.

The evidence base on hand hygiene improvement strategies is substantial, but important gaps remain for LMIC policy and implementation [4,7]. Yet, existing syntheses have tended to prioritize compliance and/or clinical endpoints without adequately relating findings to the contextual conditions that shape implementation [9]. Broader reviews and meta-analyses indicate that multimodal intervention bundles often improve compliance, though comparative effectiveness varies by component mix and study design [10].

Against this background, this review synthesizes the effectiveness of hand hygiene interventions implemented in LMIC health care facilities, including quantitative pooling where appropriate and structured effect-direction approaches where heterogeneity precludes meaningful meta-analysis. In addition, we examine contextual factors (barriers and enablers) influencing implementation and outcomes. By integrating effectiveness findings with context, this review aims to better inform implementation choices in resource-constrained settings and clarify where existing evidence is strong, where it is uncertain, and what enabling conditions appear most salient for sustained hand hygiene improvement.

## Methods

### Protocol and reporting

The review protocol was registered in the International Prospective Register of Systematic Reviews (PROSPERO; CRD420251252831) [11]. This systematic review is reported in accordance with Preferred Reporting Items for Systematic Reviews and Meta-Analyses (PRISMA) 2020 [12]; the PRISMA flow diagram is presented in Figure 1 The completed PRISMA checklist is provided as supporting information.(Supplementary file 1)

**Figure 1.**
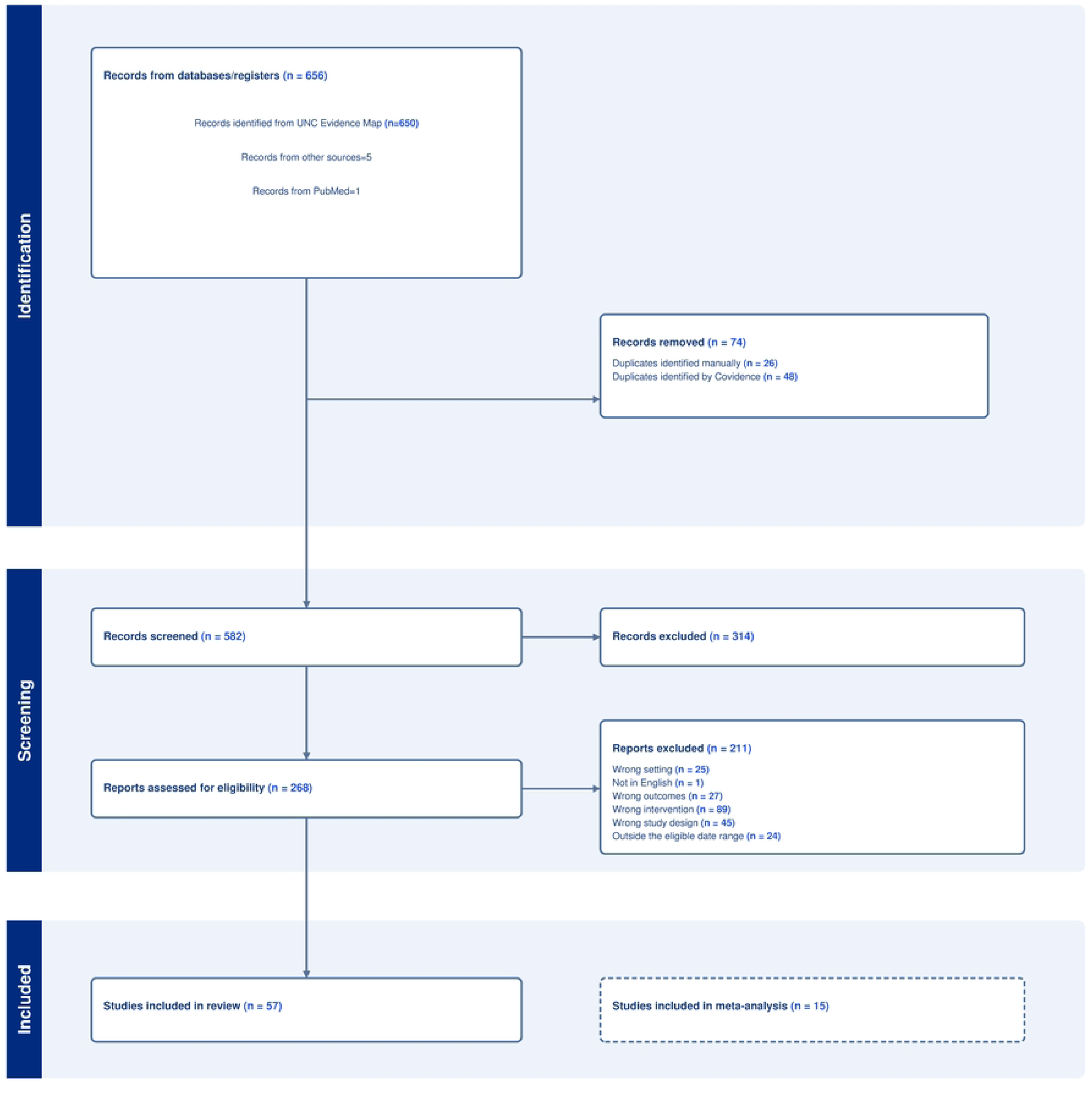
PRISMA 2020 flow diagram of study identification, screening, eligibility assessment, and inclusion.

### Search methods

This review was conducted within a broader systematic evidence map of environmental health services in healthcare facilities in low- and middle-income countries (LMICs), for which the review team was granted access to the Covidence records. The methods for the parent evidence map have been published elsewhere[13]. Briefly, the parent evidence map was developed through systematic searches of PubMed, Scopus, and Global Health (EBSCOhost) for studies on environmental health services in LMIC healthcare facilities. The original database searches were conducted on June 21, 2023, and updated on January 8, 2025. In addition, the parent evidence map included hand searches conducted in February 2025 of three journals considered highly relevant but not consistently indexed in major databases: Journal of Water and Health, Journal of Water, Sanitation, and Hygiene for Development, and H2Open Journal.

To maintain consistency with the parent evidence map, we applied the same search strategy to update the evidence-map record set for this review through November 28, 2025. Within the broader evidence-map database, we applied hand-hygiene-specific topic tags during title and abstract screening to identify potentially relevant records for subsequent full-text eligibility assessment. These tags were used to identify studies related to hand hygiene practices, hand hygiene products and infrastructure, implementation strategies, and hand hygiene outcomes.

The review was restricted to English-language studies published between January 1, 2015, and November 28, 2025. The 2015 start date was selected to align with recent global policy developments related to infection prevention and control, antimicrobial resistance, and WASH in healthcare facilities [14]. To identify additional eligible studies, we conducted backward citation chasing of relevant systematic reviews and included any eligible studies. No formal grey literature searches were conducted. Full search strings for the parent evidence map and subsequent updates are provided in the S material 1.

### Eligibility criteria

We included studies evaluating defined hand hygiene interventions implemented in healthcare facilities in low- and middle-income countries (LMICs). Studies were eligible if they: (i) were conducted in an LMIC, based on the 2022–2023 World Bank income classification [15]; (ii) were published between January 1, 2015, and November 28, 2025; (iii) were available as full-text articles in English; and (iv) were conducted in healthcare facility settings, including hospitals, health centers, clinics, wards, maternity units, outpatient departments, or other formal care settings.

Eligible interventions included strategies intended to improve hand hygiene practice, hand hygiene resources or infrastructure, or hand hygiene program implementation in healthcare facilities. Because intervention approaches were heterogeneous, detailed intervention definitions and classification rules are provided in the Supplementary Material (S S Table 4). Eligible study designs included randomized studies, quasi-experimental studies, controlled before-and-after studies, uncontrolled before-and-after evaluations, interrupted time-series studies, quality-improvement studies, mixed-methods studies, and other prospective intervention evaluations, if they evaluated a defined hand hygiene intervention and reported outcomes relevant to its effectiveness or implementation in healthcare facilities. Systematic reviews and meta-analyses were excluded from synthesis but were examined for reference mining.

### Exclusion criteria

We excluded studies conducted exclusively in non-healthcare settings, including households, schools, and other community settings. We also excluded studies focused solely on sterilization or disinfection of instruments or surgical equipment, personal hygiene practices unrelated to clinical care, household- or community-level hand hygiene, or laboratory testing of hand hygiene products without field-based implementation or outcome data. Studies that fell outside the date or language limits were also excluded ref to supplementary file 2.

### Study selection

Records from the parent evidence map and subsequent updates were imported into Covidence, where duplicates were identified and removed before screening. Two reviewers independently screened records at the title and abstract stage using Covidence. Records considered potentially eligible by either reviewer advanced to full-text screening. The same two reviewers independently assessed full texts against the predefined inclusion and exclusion criteria. Disagreements at either stage were resolved through discussion and consensus, with adjudication by a third reviewer when necessary. Reasons for exclusion at the full-text stage were recorded in Covidence [16].

### Data extraction

A data extraction file was created in Microsoft Excel, and the following information was collected: country of publication, setting of the publication, study population, country/setting, population, study design and unit of analysis, intervention components and comparator, outcome definitions and measurement methods, follow-up period, contextual factors and quantitative results (including effect estimates and measures of uncertainty where reported). One reviewer extracted data from all included studies, and a second reviewer independently verified extracted fields for accuracy and completeness; discrepancies were resolved by discussion and consensus.

### Data analysis and visualization

All analyses and figures were produced in R (version 4.5.2; R Foundation for Statistical Computing, Vienna, Austria)[17].

### Geographic distribution of included studies

A country-level choropleth map was used to summarize the geographic distribution of included studies across eligible low- and middle-income countries (LMICs) (Figure 2). Each study was linked to its study country, with multi-country studies counted once for each represented country. Study counts were aggregated at the country level and displayed using grouped categories (0, 1, 2–3, 4–6, and ≥7 studies), with numeric labels indicating the exact number of included studies per country. High-income countries were retained in grey for geographic context and excluded from the analysis, whereas LMICs with no included studies were left unshaded.

**Figure 2.**
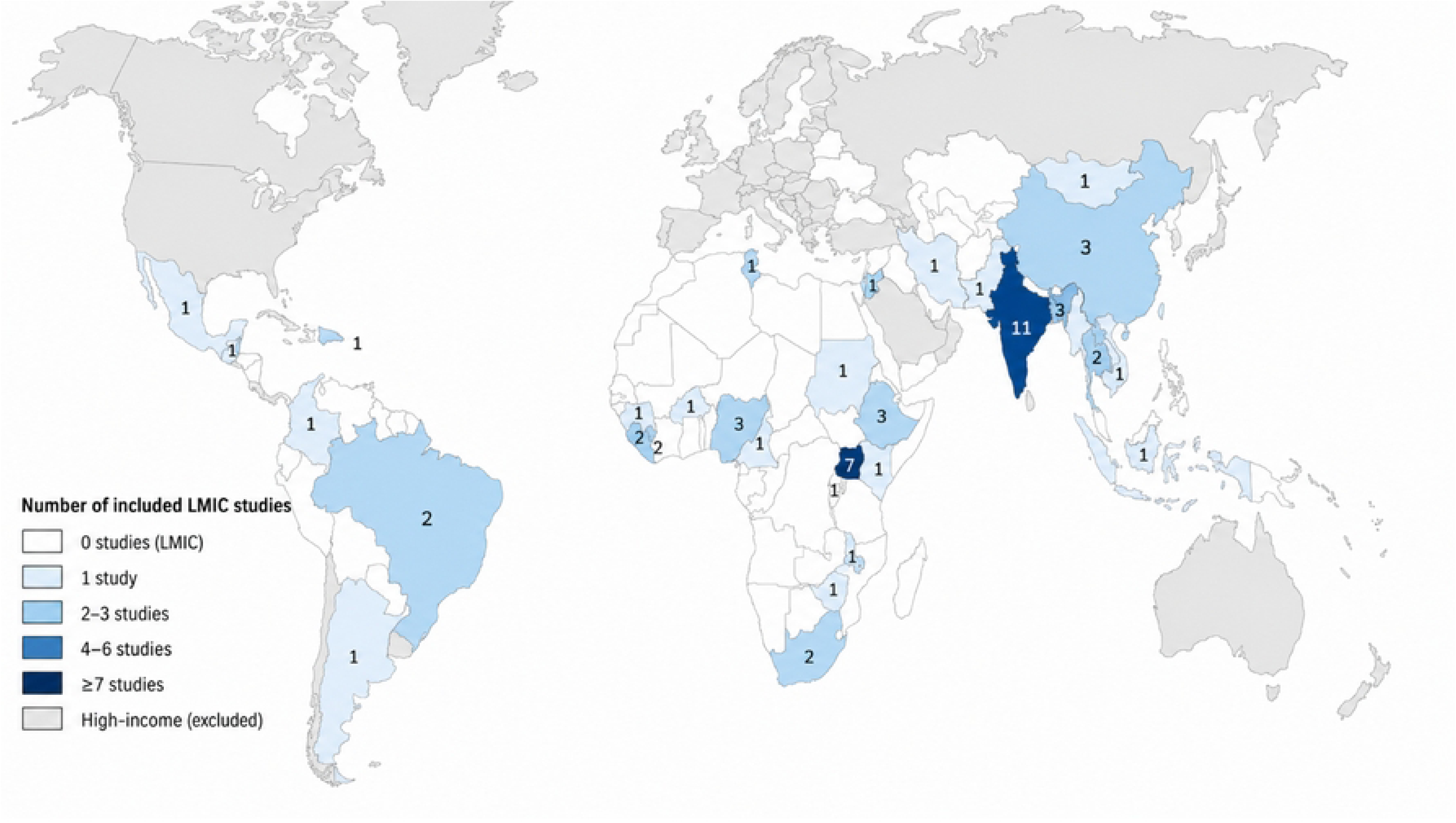
Geographic distribution of included LMIC studies.

### Study characteristics

Study characteristics were extracted using a standardized template and summarized descriptively in Table 1, with detailed study-level characteristics provided in the S S Table 3. For the summary table, we extracted publication year, WHO region, primary intervention category, outcome domains reported, and contribution to synthesis.

**Table 1.**
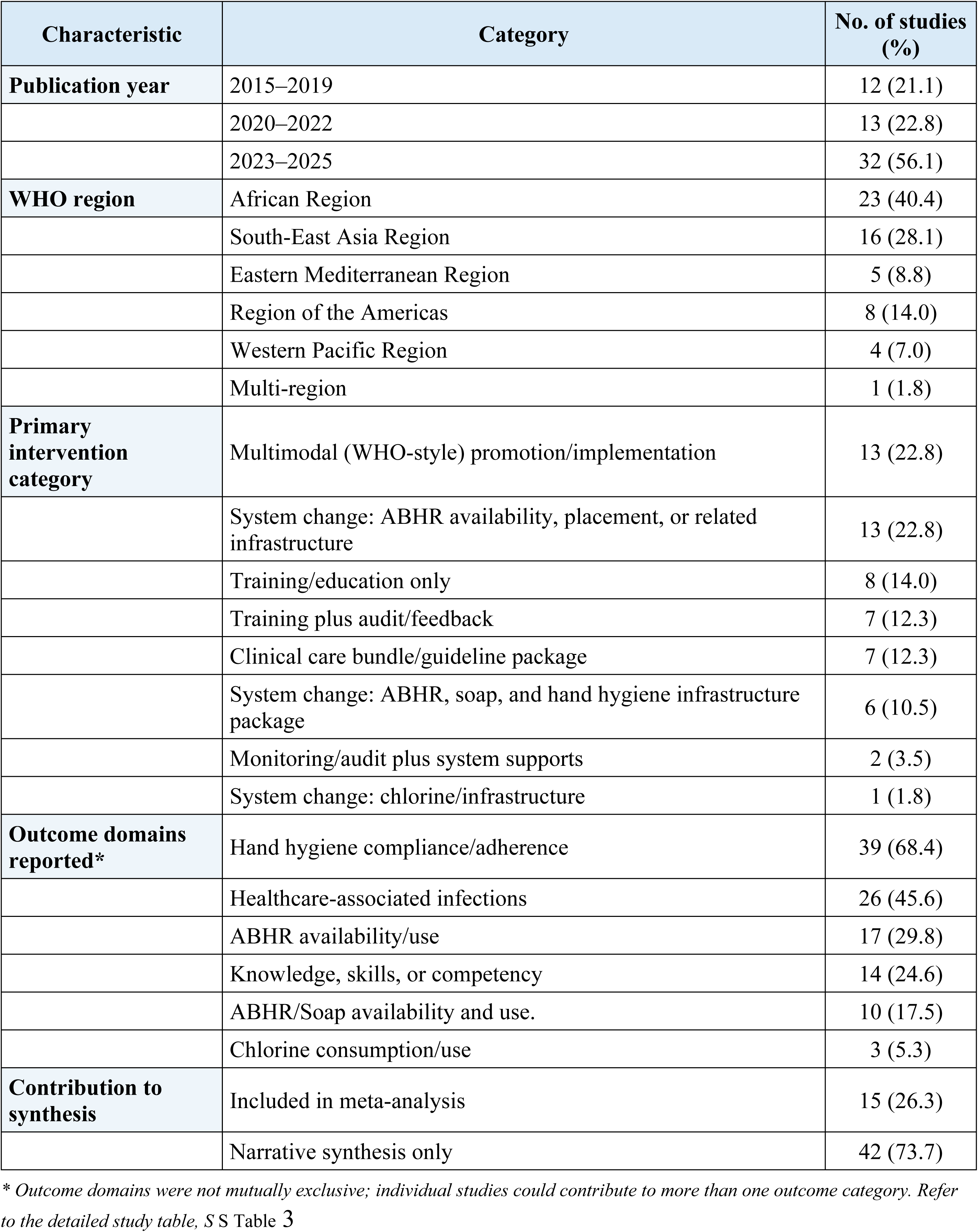
Characteristics of included studies evaluating hand hygiene interventions in low- and middle-income country health care facilities

**Table 2.** Exploratory univariable meta-regression of post-versus pre-intervention hand hygiene compliance.

| Moderator | k | Effect estimate | p-value | Residual $\tau^2$ | Pseudo-$R^2$ |
| --- | --- | --- | --- | --- | --- |
| Publication year (per 1-year increase) | 15 | RR ratio = 1.04 (95% CI 0.93 to 1.15) | 0.522 | 0.175 | 0.0% |
| Baseline compliance (per 10-percentage-point increase) | 15 | RR ratio = 0.92 (95% CI 0.82 to 1.02) | 0.108 | 0.150 | 10.5% |
| WHO African Region vs other regions | 15 | RR ratio = 1.55 (95% CI 0.95 to 2.51) | 0.079 | 0.144 | 13.5% |
| Quality score (per 10-point increase) | 15 | RR ratio = 0.99 (95% CI 0.79 to 1.25) | 0.955 | 0.190 | 0.0% |
| Intervention category overall | 15 | Omnibus p = 0.457 | 0.457 | 0.173 | 0.0% |

Intervention-family contrasts
| Contrast | k | Effect estimate | p-value |
| --- | --- | --- | --- |
| Multimodal vs other package/training | 15 | RR ratio = 1.35 (95% CI 0.78 to 2.36) | 0.282 |
| System change vs other package/training | 15 | RR ratio = 1.03 (95% CI 0.60 to 1.75) | 0.923 |

Publication year was grouped into three categories: 2015–2019, 2020–2022, and 2023–2025. Geographic distribution was summarized using WHO regions; studies conducted across more than one WHO region were classified as multi-region. Intervention categories were coded using the WHO Multimodal Hand Hygiene Improvement Strategy as an organizing framework, capturing the presence of system change, training/education, evaluation and feedback, reminders/communications, and institutional safety climate or leadership support; operational definitions and coding rules are provided in S S Table 4. Because many interventions were multi-component, each study was assigned to a single primary intervention category using prespecified decision rules based on the dominant intervention approach described by the study authors.

Reported outcomes were grouped into broad outcome domains for descriptive synthesis. These domains were hand hygiene compliance/adherence, healthcare-associated infections, ABHR availability/use, knowledge, skills, or competency, ABHR/soap availability and use, and chlorine consumption/use. Because individual studies could report more than one outcome, outcome domains were not mutually exclusive.

Studies were also categorized according to contribution to synthesis as either included in the meta-analysis or narrative synthesis only. Coding and categorization were undertaken using a piloted extraction framework, with independent checking by a second reviewer and resolution of discrepancies by consensus.

### Meta-analysis and effect-direction synthesis methods

Quantitative synthesis was undertaken for studies reporting pre- and post-intervention hand hygiene compliance with extractable numerator and denominator data, we calculated within-study risk ratios (RRs) comparing post-intervention with pre-intervention compliance and pooled these using a random-effects meta-analysis. Between-study heterogeneity was assessed using the I² statistic and τ², and a prediction interval was calculated to describe the expected range of effects in comparable settings. Exploratory univariable meta-regression was conducted to examine potential sources of heterogeneity. Publication bias was assessed using a funnel plot and Egger’s test.

For outcomes not amenable to meta-analysis, we undertook an effect-direction synthesis. Reported outcomes were grouped into broad domains, including hand hygiene compliance, knowledge, attitudes, and skills, clinical or healthcare-associated infection outcomes, alcohol-based hand rub (ABHR) availability or use, resources and infrastructure, product consumption or use, and other related outcomes. For each study–outcome pairing, the direction of effect was classified as improved, worsened, mixed, or unclear/insufficient based on the reported findings and statistical significance, where available (S table 6). These patterns were then summarized across intervention categories and outcome domains using an effect-direction plot, in which symbol color denoted effect direction and symbol shape denoted the harmonized study design class.

### Contextual moderators’ coding and qualitative synthesis

We extracted information on implementation context as contextual moderators that may influence how hand hygiene interventions were delivered, sustained and translated into outcomes. In line with our conceptual framework, contextual moderators (e.g., infrastructure, leadership, supply availability) were treated as distinct from effectiveness outcomes (e.g., hand hygiene compliance) and behavioral mediators (e.g., knowledge/attitudes/practices).

For synthesis, we summarized contextual moderators descriptively across intervention categories and domains, including studies that contributed to any quantitative pooling and those synthesized narratively, to provide a consistent cross-study picture of implementation context. We synthesized reported barriers and enablers using inductive thematic analysis, grouped them into domains, and then calculated, by intervention category, the proportion of studies reporting at least one barrier (or enabler) within each domain and visualized these proportions as heat maps (rows = intervention categories; columns = barrier/enabler domains).

### Quality assessment of included publications

Two reviewers assessed each publication’s quality using the relevant critical appraisal tool from the ICROMS framework, informed by Cochrane risk-of-bias principles[18].

## Results

### Results of search

A total of 656 records were identified (UNC Evidence Map, n = 650; PubMed, n = 1; other sources, n = 5) Figure 1. Overall, 57 studies were included in the review and 15 were included in the meta-analysis.

### Geographic distribution of included studies

Across 57 unique studies of hand hygiene interventions, evidence came from 32 LMICs Figure 2. Because four studies were conducted in more than one country, there were 61 study–country contributions in total. Study locations were concentrated in a small number of settings: India contributed 11 studies and Uganda 7. Bangladesh, China, Ethiopia, and Nigeria each contributed three studies. Brazil, Liberia, Sierra Leone, South Africa, and Thailand each contributed two studies, and the remaining 21 countries contributed one study each. Studies were reported across sub-Saharan Africa, South and East/Southeast Asia, Latin America and the Caribbean, and the Middle East.

Countries are shaded according to the number of studies included in low- and middle-income countries (LMICs), with numeric labels showing the exact study count per country. High-income countries are shown in grey for geographic context and were excluded from the analysis; LMICs without included studies are left unshaded.

### Characteristics of included studies

Among the 57 studies published between 2015 and 2025, over half (56.1%, 32/57) were published from 2023 onwards. Table 1. Characteristics of included studies evaluating hand hygiene interventions in low- and middle-income country health care facilities Most evaluations used before-and-after or quasi-experimental designs, while randomized designs were uncommon (two cluster-randomized studies). Interventions most frequently comprise multimodal (WHO-style) implementation strategies (13/57) and system change focused on alcohol-based hand rub (ABHR) availability/placement and related infrastructure (13/57). Other approaches included training/education only (8/57), training plus audit and feedback (7/57), clinical care bundles or guideline packages (7/57), and system change packages incorporating ABHR/soap and infrastructure elements (6/57); two studies evaluated monitoring/audit with system support, and one evaluated a chlorine-related system change intervention. Outcomes most assessed were hand hygiene compliance/adherence (39/57) and healthcare-associated infections (26/57). Measures of ABHR availability/use (17/57) and knowledge/skills/competency (14/57) were also frequently reported, while fewer studies assessed hand hygiene resources/infrastructure (10/57) or product consumption/use (3/57).

### Change in Hand Hygiene compliance

Figure 3 below summarizes the estimated proportion of change in HH compliance by country and study population. In a random-effects meta-analysis, post-intervention compliance was higher than pre-intervention compliance (pooled RR 1·45, 95% CI 1·20–1·74; I²=98.3%), corresponding to a 45% relative increase. Four studies showed no clear improvement or a reduction in compliance [19] 0·75 (0·64–0·87); [20] 0·87 (0·75–1·02); [21] 0·96 (0·75–1·21); [22] 0·95 (0·86–1·05), whereas three studies reported large gains [23] 2·75 (2·40–3·15); [24] 2·71 (2·47–2·97); [25] 2·62 (2·42–2·84). Based on pooled counts of reported opportunities (not accounting for clustering), crude compliance increased from 39·1% pre-intervention (6143/15711) to 60·1% post-intervention (9846/16379), an absolute increase of 21·0 percentage points, consistent with the direction of the pooled relative effect.

**Figure 3.**
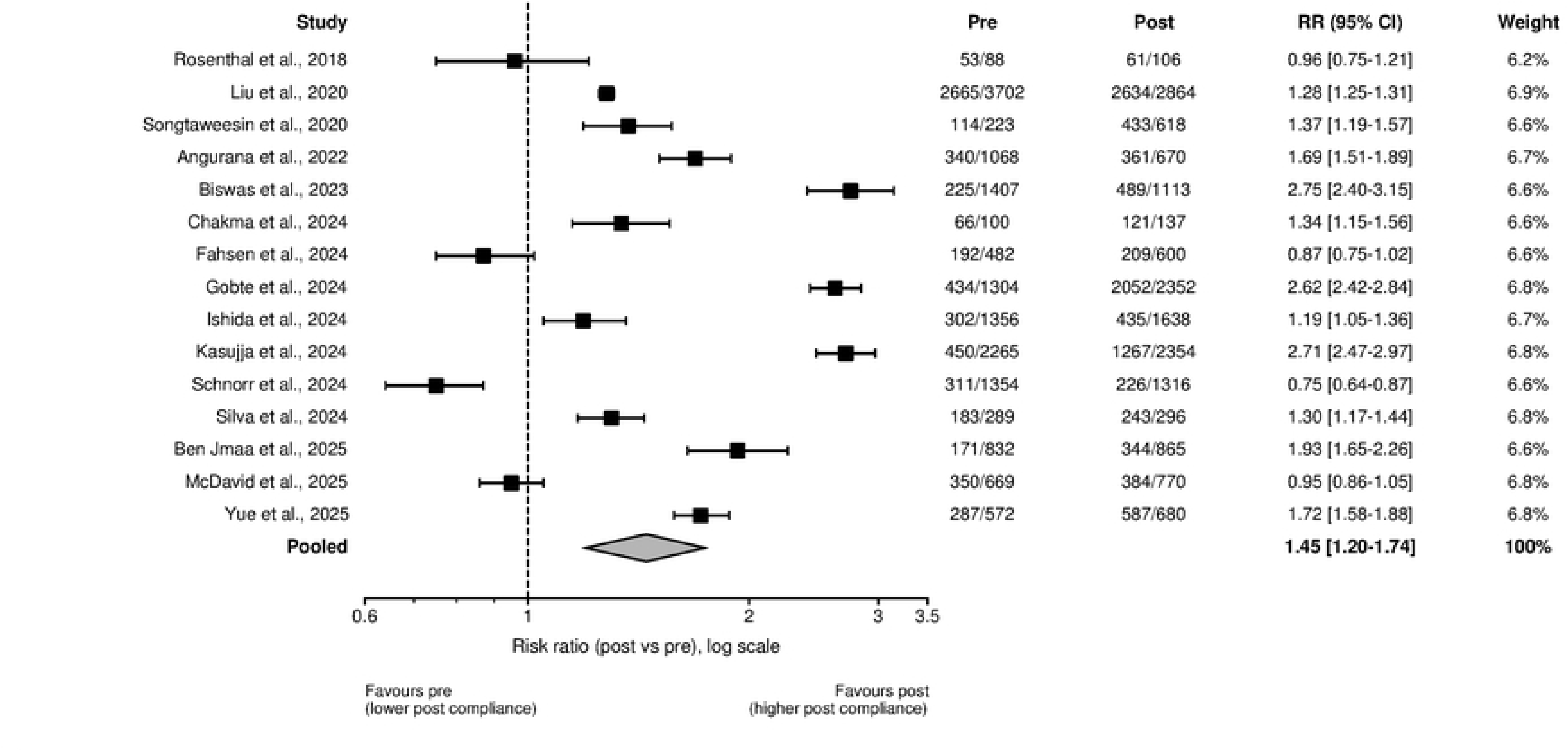
Forest plot of the change in HH compliance (n=15)

Risk ratios (RRs) compare post-intervention to pre-intervention compliance (RR>1 indicates higher post-intervention compliance). Squares are proportional to study weight and horizontal lines indicate 95% CIs; the diamond shows the pooled random effects estimate. The pooled effect was RR 1·45 (95% CI 1·20–1·74). Heterogeneity was substantial (I²=98·3%; τ²=0·126; Q (14) =826·30, p<0·001). The 95% prediction interval for the true effect in a new comparable setting was RR 0·70–2·97. Estimates are unadjusted pre–post ratios and may not account for clustering or secular trends.

### Exploratory univariable meta-regression and publication bias

Exploratory univariable meta-regression showed that publication year, study quality, and broad intervention family were not significantly associated with effect size. Studies conducted in the WHO African Region had larger relative post-intervention improvements in hand hygiene compliance than studies from other regions (RR ratio 1.55, 95% CI 1.07 to 2.23; p=0.019). Baseline compliance showed an inverse association with relative improvement (RR ratio per 10-percentage-point increase in baseline compliance, 0.92, 95% CI 0.83 to 1.01; p=0.073). Publication bias was assessed using a funnel plot and Egger’s test. Visual inspection of the funnel plot suggested slight asymmetry; however, Egger’s test did not detect statistically significant asymmetry (p=0.437; S **Error! Reference source not found.**).

### Effect-direction synthesis

Figure 4 summarizes the direction of reported effects across hand hygiene-related outcome domains, stratified by intervention category and study design. Outcomes were most frequently reported for hand hygiene compliance, knowledge/attitudes/skills, clinical or healthcare-associated infection (HAI) outcomes, and ABHR availability/use, with comparatively fewer observations for resources/infrastructure, product consumption/use, and other outcomes. Across all intervention categories represented (training-based interventions, monitoring/audit approaches, multimodal WHO-style strategies, system change interventions, and clinical care bundles/guideline packages), most reported effects indicated improvement for hand hygiene compliance and for behavioral and system-related domains. Mixed and unclear findings were also observed across multiple domains, and were most prominent for clinical/HAI outcomes, where results were more heterogeneous. Worsened effect directions were reported infrequently. Across the effect-direction synthesis, 79% of reported study–outcome effects were favorable (77/97). Improvements were more consistently positive for hand hygiene compliance and supply/use measures than for healthcare-associated infection outcomes, which were less frequently reported and more often showed no clear change; worsened directions were rare (3/97).

**Figure 4.**
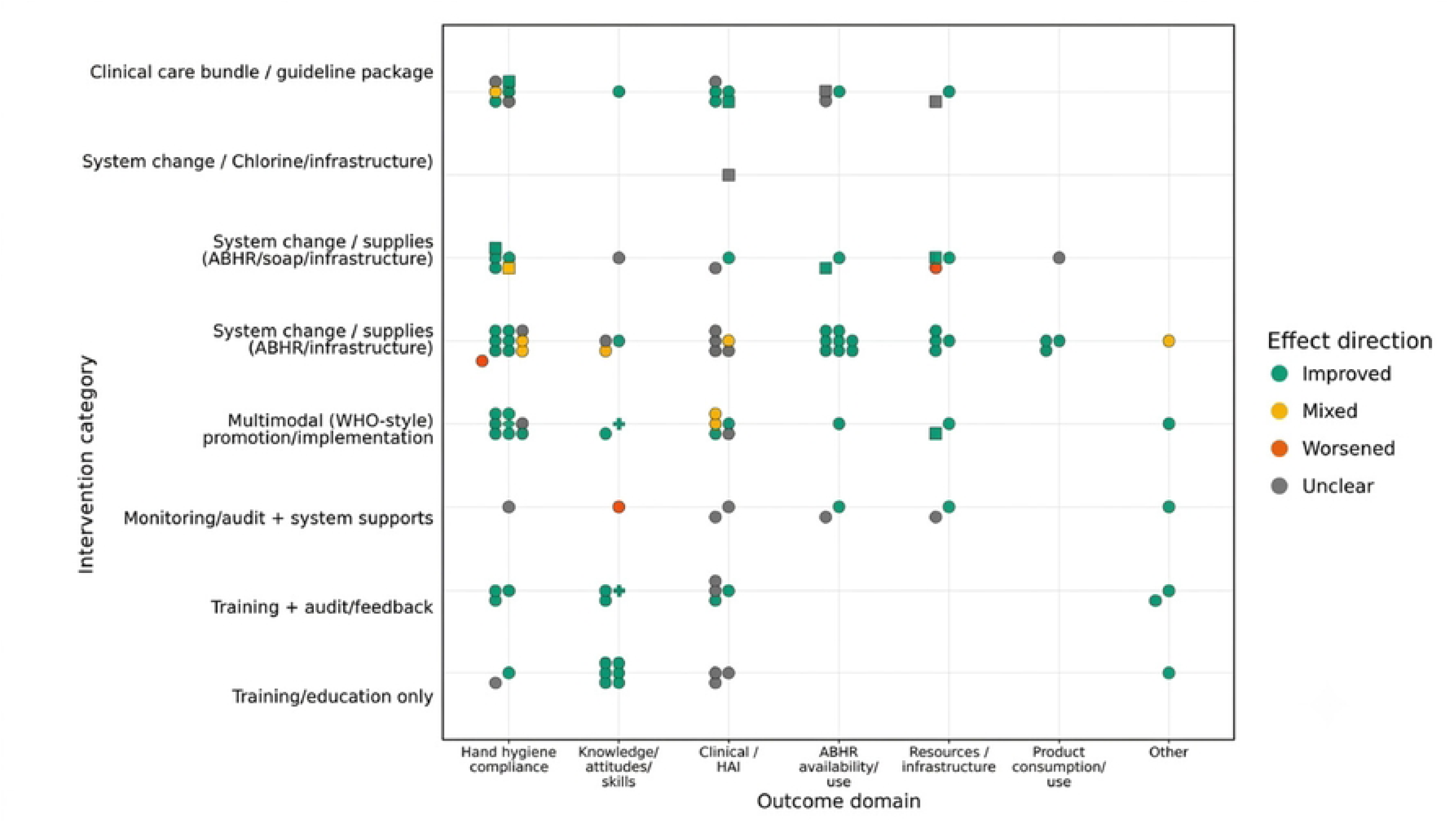
Effect direction plot of hand hygiene interventions by intervention category and outcome domain.

Each symbol represents an individual study outcome and summarizes the reported direction of effect as improved, mixed, worsened, or unclear. Symbol color indicates effect direction, and symbol shape indicates the harmonized study design class: circles denote non-randomized interventional, before–and–after, quasi-experimental, and quality-improvement studies; squares denote mixed-methods, observational, implementation case-study, or other non-randomized designs; and signs denote randomized studies. The figure illustrates the predominance of positive effects for behavioral and system-related outcomes.

### Contextual barriers to implementation

**Figure 5** summarizes barriers across nine domains as the proportion of studies within each intervention category reporting ≥1 barrier in that domain. Across intervention types, barriers were most frequently reported in audit/data systems (ranging from 1/6 to 4/6 and up to 4/11), supply shortages (up to 6/11 and 3/6), and staffing/workload/time pressure (up to 4/6). Audit/data barriers included difficulties conducting or sustaining hand-hygiene observations, incomplete or inconsistent documentation, and challenges using audit results for feedback and improvement [26–28]. Supply barriers reflected inadequate or inconsistent access to supplies such as ABHR [29–31], while staffing/workload barriers were described as time pressure and workload constraints limiting either compliance or implementation activities [19,32,33]. Barriers related to WASH/infrastructure and implementation support were also commonly reported, including constraints in water access, sinks, soap, or handwashing facilities particularly among studies evaluating system change interventions that incorporated ABHR, soap, and infrastructure components (5/6 studies). [29,34–36] and limitations in supervision, coordination, or operational support needed to sustain implementation processes (e.g., 4/6 in training + audit/feedback; 3/11 in ABHR/infrastructure system change) [28,37–39]. In contrast, leadership/governance and financing constraints were less frequently reported overall (often 0/6–3/11 for leadership/governance and 0/6–2/11 for financing), though present in a minority of studies [40,41].

**Figure 5.**
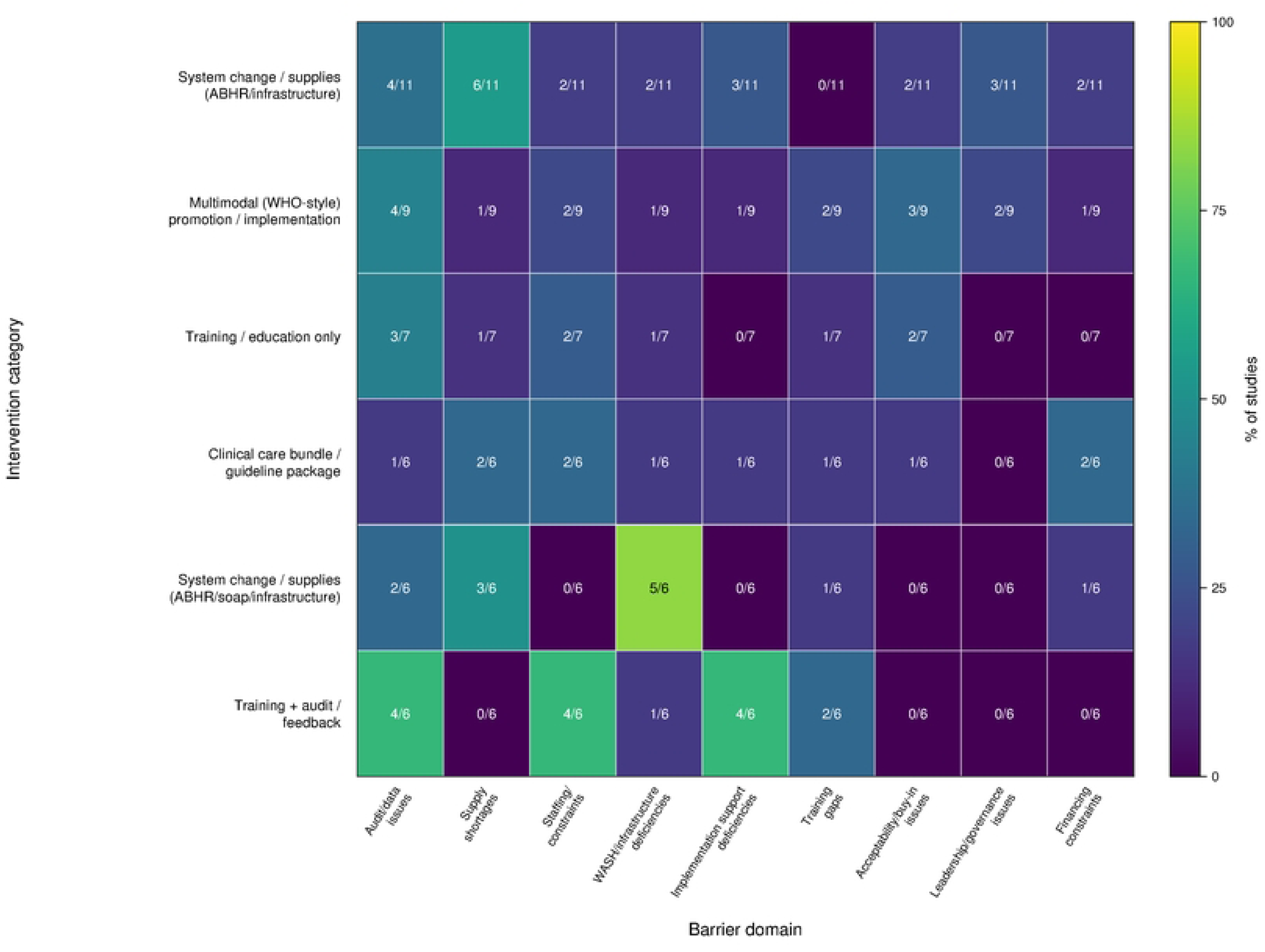
Implementation barrier domains by intervention category. Heat map showing the percentage of studies within each primary intervention category that reported ≥1 implementation barrier in each barrier domain

Barrier profiles varied by intervention category. In system change focused on ABHR/infrastructure, supply shortages (6/11) and audit/data issues (4/11) were most frequently reported (Figure 5), with study descriptions citing ABHR availability and monitoring/measurement challenges [25,30,42] and audit/feedback data-system limitations[23,43]. In system change incorporating ABHR/soap/infrastructure, WASH/infrastructure constraints predominated (5/6) (e.g., water, soap, handwashing facilities), while other domains were reported less consistently [29,34–36]. Training + audit/feedback interventions most often reported co-occurring barriers in audit/data (4/6), staffing/time pressure (4/6), and implementation support (4/6), including challenges maintaining observation/feedback processes, staff time to deliver audit cycles, and the coordination required to sustain them [28,37–39]. In multimodal (WHO-style) interventions, barriers were distributed across domains, commonly including audit/data issues (4/9) [26,44,45] and, in some studies, acceptability/buy-in (3/9) such as limited staff engagement with monitoring processes or reluctance to consistently adopt intervention components [46]. Where reported, financing constraints were described as budget or procurement limitations affecting sustainability (e.g., sourcing inputs for ABHR production or maintaining components over time) [26,41], and leadership/governance issues included challenges maintaining program oversight and continuity under leadership change, reliance on champions, and knowledge transfer under staff turnover or rotation [40,46]. Study-level descriptions of the specific barriers coded within each domain and any barriers not captured in **Figure 5** are provided in S S Table 1.

### Contextual enablers by intervention category

**Figure 6** summarizes contextual enablers across eight domains, expressed as the percentage of studies within each intervention category reporting ≥1 enabler in that domain (denominators shown per category). Across categories, enablers were most frequently reported for audit/data and training. Audit/data enablers included structured compliance monitoring (e.g., direct observation or audit tools), routine availability of compliance data, and mechanisms to feed results back to staff (e.g., feedback meetings or reporting structures) [21,47,48]. Training enablers included delivery of formal training and refresher sessions, bedside coaching/mentorship, and cascade training approaches where trained staff trained others [23,33]. Leadership/governance and supplies/equipment enablers were also reported in specific intervention types, including leadership sponsorship and hand hygiene champion structures [24,40,46]and reliable availability or placement of ABHR/hand-hygiene materials and related equipment (e.g., point-of-care ABHR provision or strengthened stock management) [30,41]. In contrast, acceptability/buy-in and WASH/infrastructure enablers were reported infrequently across categories (generally 0/6–1/10 for WASH/infrastructure and 0/7–2/10 for acceptability/buy-in. Where present, acceptability enablers were described as staff engagement or positive perceptions of intervention components [45,49], while WASH/infrastructure enablers were described as supportive physical conditions such as functioning handwashing stations or sink-based cues[20,37].

**Figure 6.**
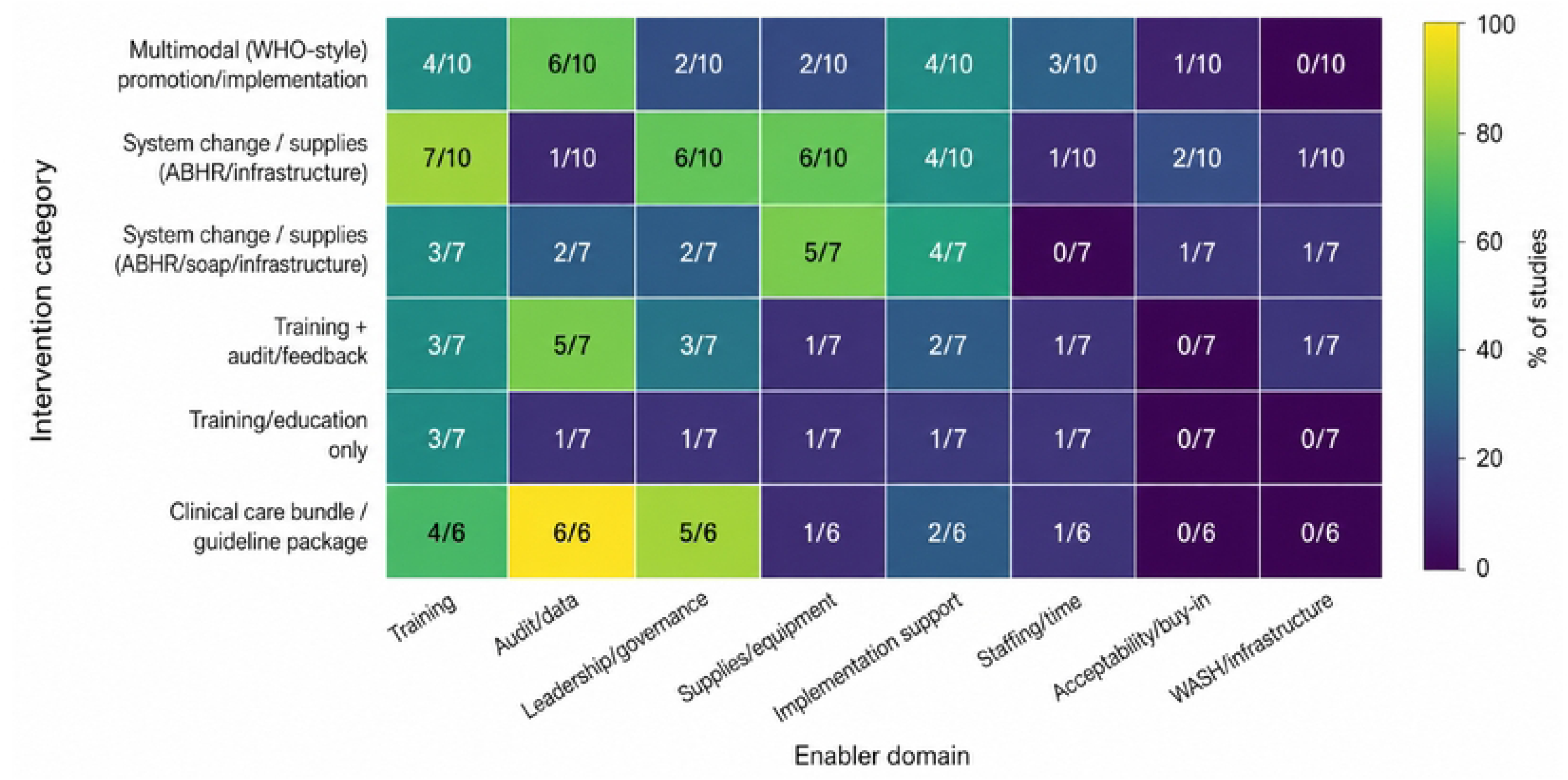
Contextual enabler domains by intervention category. Cell labels indicate k/n (number of studies reporting an enabler in that domain / total studies contributing enabler information for that intervention category).

Enabler profiles varied by intervention category. Multimodal (WHO-style) interventions (n=10) most often reported Audit/data (6/10), with training and implementation support each reported in 4/10 studies [44,50]. In system change focused on ABHR/infrastructure (n=10), enablers were most frequently reported for Training (7/10), leadership/governance (6/10), and supplies/equipment (6/10), while audit/data enablers were reported less often (1/10) [27,80]. System change incorporating ABHR/soap/infrastructure (n=7) most often reported supplies/equipment (5/7) and implementation support (4/7) [34,36,40]. Training + audit/feedback interventions (n=7) most frequently reported audit/data (5/7), with studies describing structured observation and feedback processes supported by trained IPC personnel and reporting/feedback routines [28,37,39,48]. Training/education-only studies (n=7) reported enablers less consistently across domains (typically 1/7 outside training) [51,52]. Clinical care bundle/guideline package studies (n=6) reported the most consistent enabling conditions, particularly audit/data (6/6) and leadership/governance (5/6), alongside training (4/6) [31,53].

Study-level descriptions and coding for each domain are provided in S S Table 2.

### Study-level methodological appraisal

Methodological quality judgments varied across domains (S S figure 2). Exposure had the highest proportion of low-risk/good ratings (66%), followed by contextual rigor (53%). Allocation had the highest proportion of high-risk/poor ratings (54%), with confounding also frequently rated high risk/poor (41%). Several domains were predominantly rated unclear/some concerns, including selection (86%), follow-up (81%), outcome (79%), and fidelity & sustainability (72%). Overall methodological judgments were most commonly unclear/some concerns (87%), with fewer rated low risk/good (11%) and very few rated high risk/poor (2%).

## Discussion

This review synthesized evidence from 57 studies across 32 low- and middle-income countries (LMICs) and found that hand hygiene interventions were generally associated with improved observed hand hygiene compliance, although effects varied substantially across settings and study designs. Taken together, the evidence suggests that hand hygiene interventions may improve compliance, although their effects appear to vary considerably across settings. This variability is consistent with the exploratory meta-regression, which indicated larger relative improvements in studies from the WHO African Region and a possible inverse association between baseline compliance and relative improvement. It is further supported by the contextual synthesis, in which implementation was frequently constrained by weaknesses in audit and data systems, supply shortages, staffing and workload pressures, and WASH infrastructure, whereas stronger performance was more often reported in the presence of structured monitoring and feedback, ongoing training, leadership support, and reliable point-of-care access to hand hygiene materials. These findings suggest that intervention effectiveness is shaped not only by the intervention components themselves but also by the broader implementation environment. Although formal assessment of these factors was limited by inconsistent reporting and small numbers of studies within comparable categories, contextual differences likely contributed importantly to the observed heterogeneity. In contrast, evidence for consistent reductions in healthcare-associated infections (HAIs) remained mixed/uncertain, consistent with the more heterogeneous direction of effects reported for clinical/HAI outcomes across the broader effect-direction synthesis. This divergence is plausible: observed compliance is proximal to intervention delivery and can respond rapidly to improved access and reinforcement, whereas HAIs are influenced by multiple concurrent infection prevention and control (IPC) practices, case-mix, surveillance sensitivity, and follow-up duration [6,9,10].

Within the effect-direction synthesis multimodal World Health Organization (WHO)-style strategies and system change/supplies interventions showed predominantly favorable directions for observed hand hygiene compliance. For multimodal strategies, most classifiable studies showed positive effects, with only one study reporting an unclear effect direction. For system change/supplies interventions, most studies also showed positive effects, although a small number reported mixed or negative findings. These intervention packages often combined education with cues and leadership-led promotion, such as mobile or video reminders, posters, portable alcohol-based hand rub (ABHR), recurring feedback cycles, and quality improvement activities [24,44,45]. System change/supplies approaches emphasized strengthening point-of-care access to ABHR (e.g., dispenser placement, portable bottles/holsters, and local production and distribution) and were also associated with some of the larger pre-post improvements where numerator-denominator data were available.

Illustrative system-focused models included local ABHR production with ward-level dispenser mounting and quality control [19], wearable point-of-care ABHR to improve workflow [54], and centralized ABHR production/distribution approaches [41,42].

Training was rarely a uniform, stand-alone component. Instead, studies implemented training packages with different delivery modes and varying levels of ongoing support, including self-paced digital training (e.g., a modular online IPC course that included a dedicated hand hygiene module) [28], live online WHO-based modules [52], and in-person approaches with implementation support (e.g., workshops, demonstrations, and reinforcement through reminders and feedback structures) [44]. A smaller subset tested simulation-based approaches, including virtual reality (VR) training, delivered as repeated short practice sessions with real-time feedback [55]. Across modalities, the differentiating feature was not the platform but whether training was embedded within opportunity (reliable access to products and workflow fit) and reinforcement (monitoring/feedback routines, cues, and supervision), rather than delivered as a one-off educational session.

Implementation findings help interpret the observed heterogeneity and the weaker, more variable signals for HAIs. Across intervention categories, frequently reported barriers included limitations in audit/data systems, supply shortages, and staffing constraints and workload/time pressure, with additional constraints related to water, sanitation, and hygiene (WASH) infrastructure and implementation support. These barriers represent practical bottlenecks in delivery and sustainment, including difficulty maintaining observations and documentation, gaps in ABHR and other consumables, time pressure limiting practice or program activities, and infrastructure constraints (water, soap, sinks/handwashing facilities) that reduce feasibility at the point of care. On the other hand, implementation success was often supported by structured monitoring and feedback (routine availability and use of compliance data) and ongoing training supports (refreshers, coaching/mentorship, cascade training), with leadership/governance and supplies/equipment enablers prominent in some categories. Together, these patterns suggest a pathway in which skills and expectations are strengthened through training, opportunity is enabled through reliable point-of-care resources and functional infrastructure, and improvements are sustained through feedback, supervision, and governance structures [4]. These findings have important implications for policy and implementation. Hand hygiene improvement in LMIC health-care facilities should be treated as a health-systems intervention rather than a short-term educational activity. National and subnational programs should define minimum point-of-care hand hygiene service standards, including availability, placement, replenishment, and maintenance of ABHR, soap, water, and hand hygiene stations. These standards should be linked to procurement systems, facility budgets, and accountability mechanisms, with responsibility clearly assigned to facility IPC teams, supply-chain staff, and district or subnational health managers. Routine indicators, such as point-of-care ABHR availability and stockout frequency, could provide practical measures for monitoring implementation gaps.

At facility level, one-off distribution of hand hygiene materials is unlikely to be sufficient. Sustained improvement requires continuity mechanisms that keep hand hygiene resources functional during routine care, including procurement planning, buffer stocks, refill workflows, dispenser maintenance, stock monitoring, and rapid troubleshooting when supplies are interrupted. Where local ABHR production is used, quality control, reliable input sourcing, and distribution arrangements should be incorporated into routine facility operations. ABHR access should also be paired with maintenance of soap-and-water infrastructure where indicated, because alcohol-based hand rub and handwashing facilities are complementary requirements in many care contexts. Routine functionality checks for sinks, water, soap, dispensers, and ABHR stocks, with documented corrective actions, could help translate system-change interventions into sustained point-of-care availability.

The implementation findings also suggest that training is most likely to be effective when embedded within a broader reinforcement package. Training modalities can be adapted to local context, including digital modules, live online sessions, in-person coaching, mentorship, and simulation-based approaches where feasible. However, training should be paired with reminders or cues, feasible low-burden monitoring, regular feedback, and designated hand hygiene champions. Protecting time for IPC functions, formalizing succession planning for champions, and integrating hand hygiene into onboarding and refresher systems could help reduce the effects of staff turnover and workload pressure. Assigning responsibility for regular feedback reporting and action tracking to unit managers and facility IPC leads could also strengthen the link between audit data and practical improvement.

Study quality appraisal supports cautious interpretation, particularly for clinical endpoints. Overall methodological judgments were most classified as unclear or as having some concerns, with fewer studies rated as low risk/good quality and very few rated as high risk/poor quality.

At the domain level, allocation and confounding were most often rated high risk/poor, while selection, follow-up, outcome assessment, and fidelity/sustainability were commonly unclear. This profile is consistent with literature dominated by before-after and quasi-experimental designs in which concurrent IPC changes, secular trends, clustering, and measurement reactivity can bias effect estimates, most directly for observed compliance and even more so for HAI outcomes that depend on surveillance capacity and consistent definitions [56]. Overall, the evidence strongly supports directional improvement in observed compliance, while uncertainty remains about the magnitude and durability of effects and about consistent translation into HAI reductions across settings.

Several limitations should be considered when interpreting these findings. Only 15 studies contributed to the pooled analysis; therefore, meta-regression was exploratory and restricted to univariable models. Multivariable meta-regression was not undertaken because the small number of pooled studies and sparse distribution of candidate moderators would have produced underpowered and unstable models. In addition, most included studies used before–after, quasi-experimental, or related non-randomized designs, making it difficult to separate intervention effects from secular trends, concurrent IPC activities, measurement reactivity, or changes in surveillance practice. Reporting of implementation context, intervention fidelity, and sustainability was also inconsistent, limiting our ability to compare which intervention components were most important across settings.

Future research should move beyond short-term compliance outcomes and evaluate implementation strategies under routine health-system conditions. Pragmatic studies are needed to compare training delivery modalities, including online training, in-person coaching, mentorship, and simulation-based approaches such as virtual reality, within broader implementation packages. Future evaluations should also include cost, feasibility, reach, sustainability, and equity outcomes, including whether interventions perform differently by facility level, baseline infrastructure, staffing constraints, and supply-chain capacity. Studies linking hand hygiene improvements to patient-level HAI outcomes, while accounting for contextual and implementation factors, would provide stronger evidence for scale-up and policy investment.

## Conclusion

Hand hygiene interventions in LMIC healthcare facilities were generally associated with improved observed compliance, but effects varied substantially and evidence for consistent reductions in healthcare-associated infections remained mixed. The findings indicate that sustained improvement depends not only on education or reminders, but on the systems that enable hand hygiene at the point of care, including reliable supplies, functional WASH infrastructure, monitoring and feedback, leadership support, and protected implementation time. Hand hygiene should therefore be implemented as a health-systems intervention embedded within IPC and WASH programs, rather than as a stand-alone behavior-change activity. Future research should assess sustainability, cost, equity, and whether improvements in compliance translate into reductions in patient-level infection outcomes.

## Data Availability

All data underlying the findings of this manuscript are available within the manuscript and its Supporting Information files.

## Ethical Considerations

This review utilized data derived exclusively from published, peer-reviewed sources and therefore did not require ethical approval. Nonetheless, the research team adhered to established standards of scholarly integrity, ensuring accurate citation, transparent reporting, and acknowledgement of all original authors.

## Data sharing statement

This systematic review did not collect individual participant data. The dataset consists of extracted study-level data from published articles, including study characteristics, intervention details, outcome domains, effect-direction coding, implementation barriers and enablers, quality appraisal results, and extracted numerator and denominator data used for the meta-analysis of hand hygiene compliance.

The review protocol is available in PROSPERO (CRD420251252831). Search strategies, study characteristics, intervention classification rules, coding frameworks, effect-direction coding, quality appraisal summaries, and supplementary tables are provided in the manuscript and supplementary materials.

## Supplementary Materials

### S material 1. Search String

#### Healthcare facilities

healthcare OR “health care” OR hospital$ OR clinic$ OR “health facilit*” OR “health center$” OR healthcenter$ OR healthcentre$ OR “health centre$” OR “health post$” OR healthpost$ OR “health setting*” OR “medical facilit*” OR “medical center$” OR “medical centre$” OR “medical post$” OR “medical setting*” OR “delivery facilit*” OR “delivery center$” OR “delivery centre$” OR “delivery clinic$” OR “birth facilit*” OR “birth center$” OR “birth centre$” OR “birth clinic$” OR “matern* facilit*” OR “matern* center$” OR “matern* centre$” OR “matern* clinic$” OR dispensary OR dispensaries

#### Low- and middle-income countries

Afghanistan OR Algeria OR Angola OR Anguilla OR Antigua OR Barbuda OR Argentina OR Armenia OR Armenian OR Aruba OR Azerbaijan OR Bahamas OR Bahrain OR Bangladesh OR Barbados OR Benin OR Byelarus OR Byelorussian OR Belarus OR Belorussian OR Belorussia OR Belize OR Bhutan OR Bolivia OR Botswana OR Brazil OR Brunei OR “Burkina Faso” OR “Burkina Fasso” OR “Upper Volta” OR Burundi OR Urundi OR Cambodia OR “Khmer Republic” OR Kampuchea OR Cameroon OR Cameroons OR Cameron OR Camerons OR “Cape Verde” OR “Cayman Islands” OR “Central African Republic” OR Chad OR Chile OR China OR Colombia OR Comoros OR “Comoro Islands” OR Comores OR Mayotte OR Congo OR Zaire OR “Cook Islands” OR “Costa Rica” OR “Cote d’Ivoire” OR “Ivory Coast” OR Croatia OR Cuba OR Cyprus OR Djibouti OR “French Somaliland” OR Dominica OR “Dominican Republic” OR “East Timor” OR “East Timur” OR “Timor Leste” OR Ecuador OR Egypt OR “United Arab Republic” OR “El Salvador” OR Eritrea OR Ethiopia OR “Falkland Islands” OR “Las Malvinas” OR Fiji OR Gabon OR “Gabonese Republic” OR Gambia OR Gaza OR “Georgia Republic” OR “Georgian Republic” OR Ghana OR “Gold Coast” OR Greece OR Grenada OR Guatemala OR Guinea OR Guam OR Guadeloupe OR Guiana OR Guyana OR Haiti OR Honduras OR “Hong Kong” OR India OR Maldives OR Indonesia OR Iran OR Iraq OR Jamaica OR Jordan OR Kazakhstan OR Kazakh OR Kenya OR Kiribati OR Korea OR Kosovo OR Kuwait OR Kyrgyzstan OR Kirghizia OR “Kyrgyz Republic” OR Kirghiz OR Kirgizstan OR “Lao PDR” OR Laos OR Lebanon OR Lesotho OR Basutoland OR Liberia OR Libya OR Macau OR Madagascar OR “Malagasy Republic” OR Maldives OR Malaysia OR Malaya OR Malay OR Sabah OR Sarawak OR Malawi OR Nyasaland OR Mali OR Malta OR “Marshall Islands” OR Martinique OR Mauritania OR Mauritius OR “Agalega Islands” OR Mexico OR Micronesia OR “Middle East” OR Mongolia OR Montserrat OR Morocco OR Ifni OR Mozambique OR Myanmar OR Myanma OR Burma OR Namibia OR Nauru OR Nepal OR Niui OR “Netherlands Antilles” OR “New Caledonia” OR Nicaragua OR Niger OR Nigeria OR “Northern Mariana Islands” OR Oman OR Mayotte OR Muscat OR Pakistan OR Palau OR Palestine OR Panama OR Paraguay OR Peru OR Philippines OR Philipines OR Phillipines OR Phillippines OR Polynesia OR “Puerto Rico” OR Qatar OR Reunion OR Rwanda OR Ruanda OR “Saint Kitts” OR “St Kitts” OR Nevis OR “Saint Lucia” OR “St Lucia” OR “Saint Vincent” OR “St Vincent” OR Grenadines OR Samoa OR “Samoan Islands” OR “Navigator Island” OR “Navigator Islands” OR “Sao Tome” OR “Saudi Arabia” OR Senegal OR Serbia OR Montenegro OR Seychelles OR “Sierra Leone” OR Singapore OR “Sri Lanka” OR Ceylon OR “Solomon Islands” OR Somalia OR “South Africa” OR Sudan OR Suriname OR Surinam OR Swaziland OR Syria OR Tajikistan OR Tadzhikistan OR Tadjikistan OR Tadzhik OR Tanzania OR Thailand OR Togo OR “Togolese Republic” OR Tokelau OR Tonga OR Trinidad OR Tobago OR Tunisia OR Turkey OR Turkmenistan OR Turkmen OR “Turks Caicos” OR “Turks and Caicos” OR Tuvalu OR Uganda OR “United Arab Emirates” OR Uruguay OR Uzbekistan OR Uzbek OR Vanuatu OR “New Hebrides” OR Venezuela OR Vietnam OR “Viet Nam” OR “Virgin Islands” OR “West Bank” OR Yemen OR Yugoslavia OR Zambia OR Zimbabwe

---

**S Table 1.**
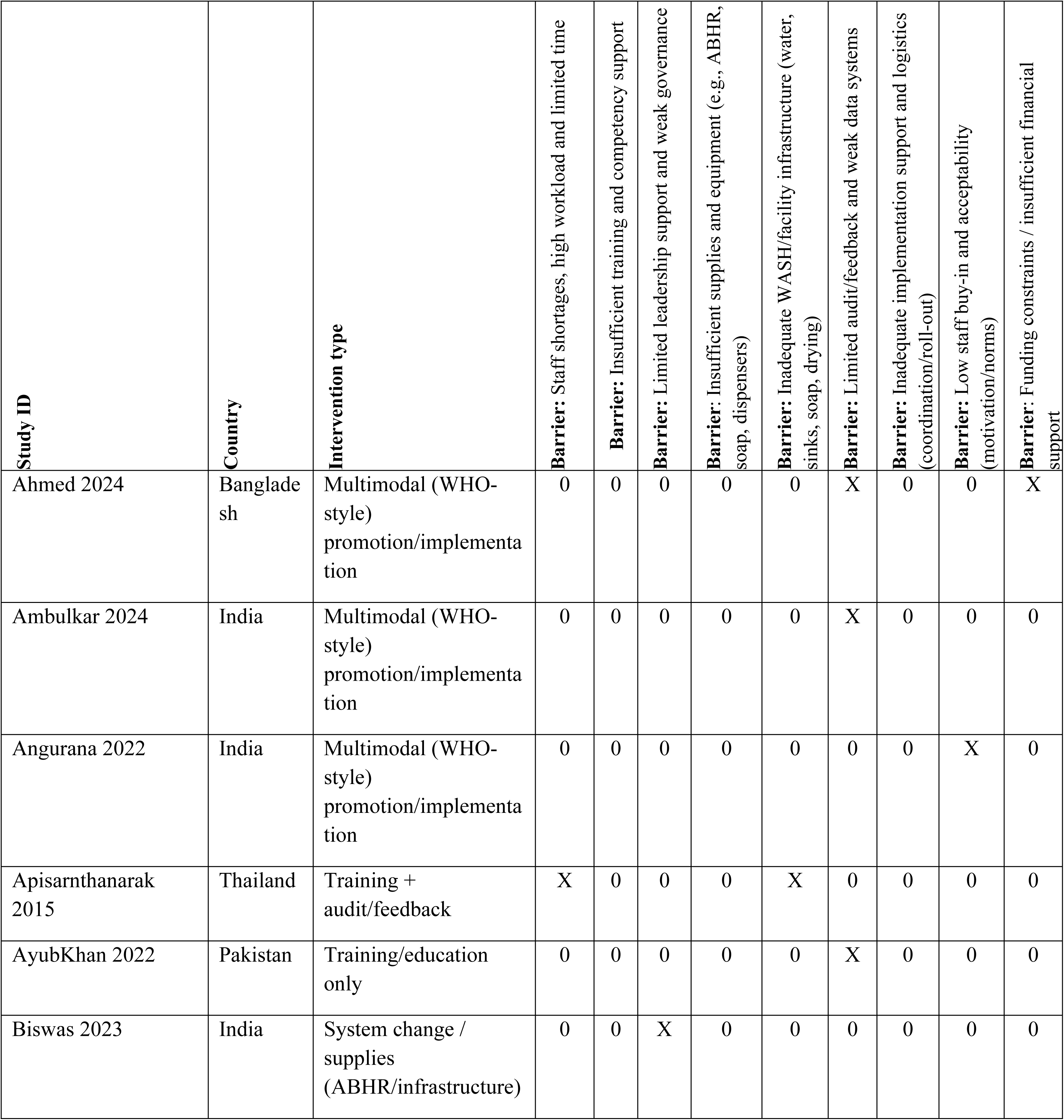
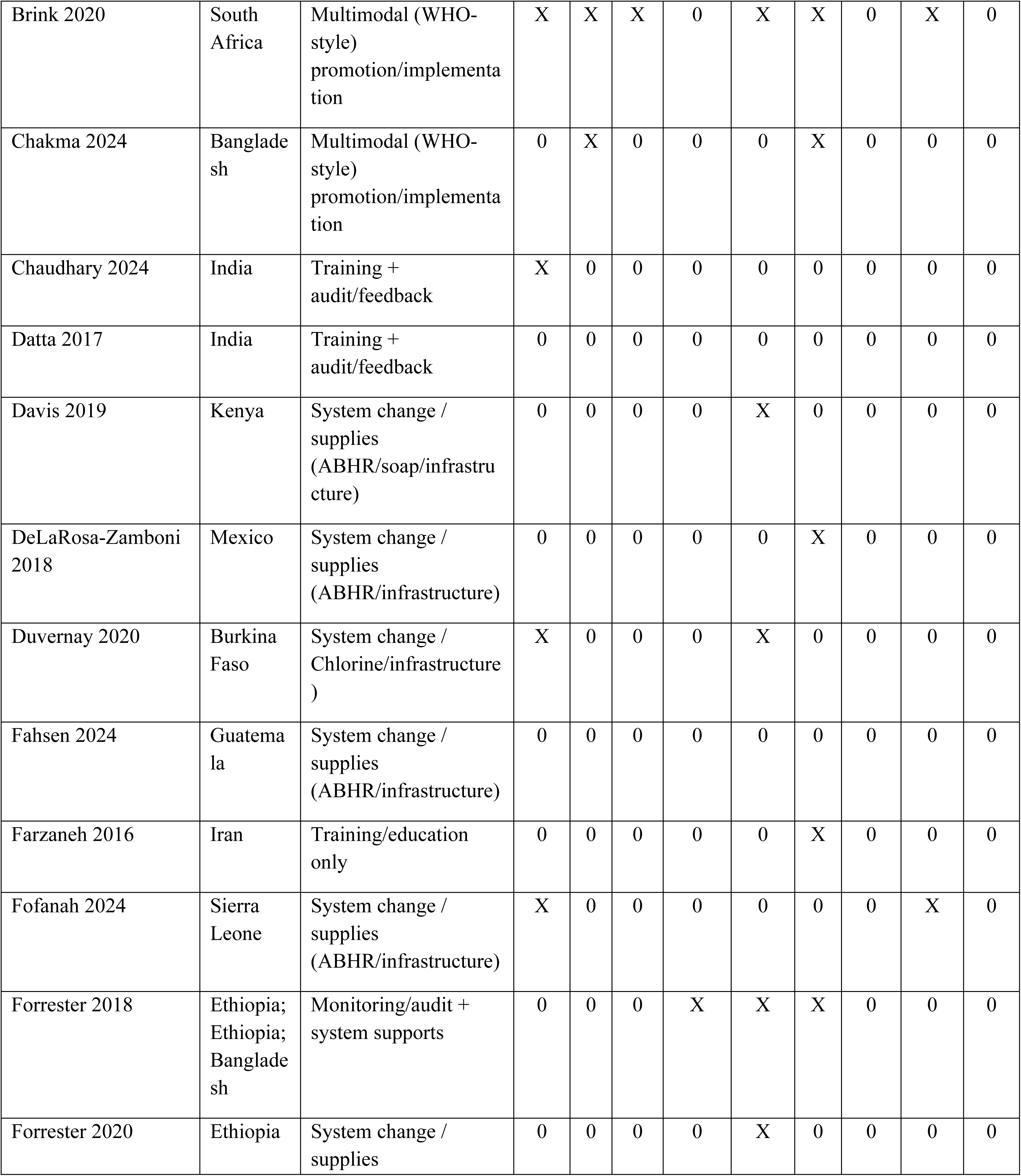
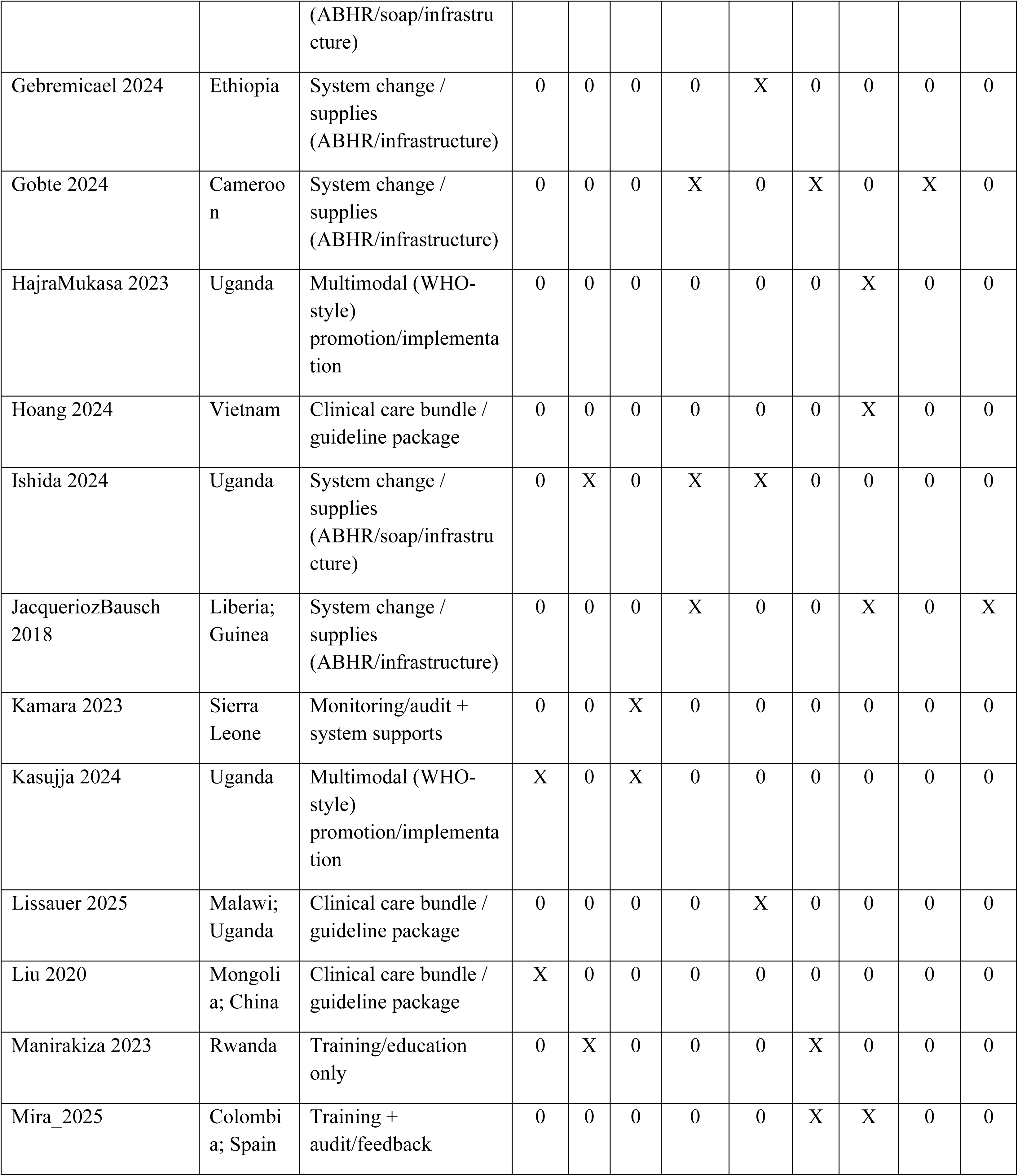
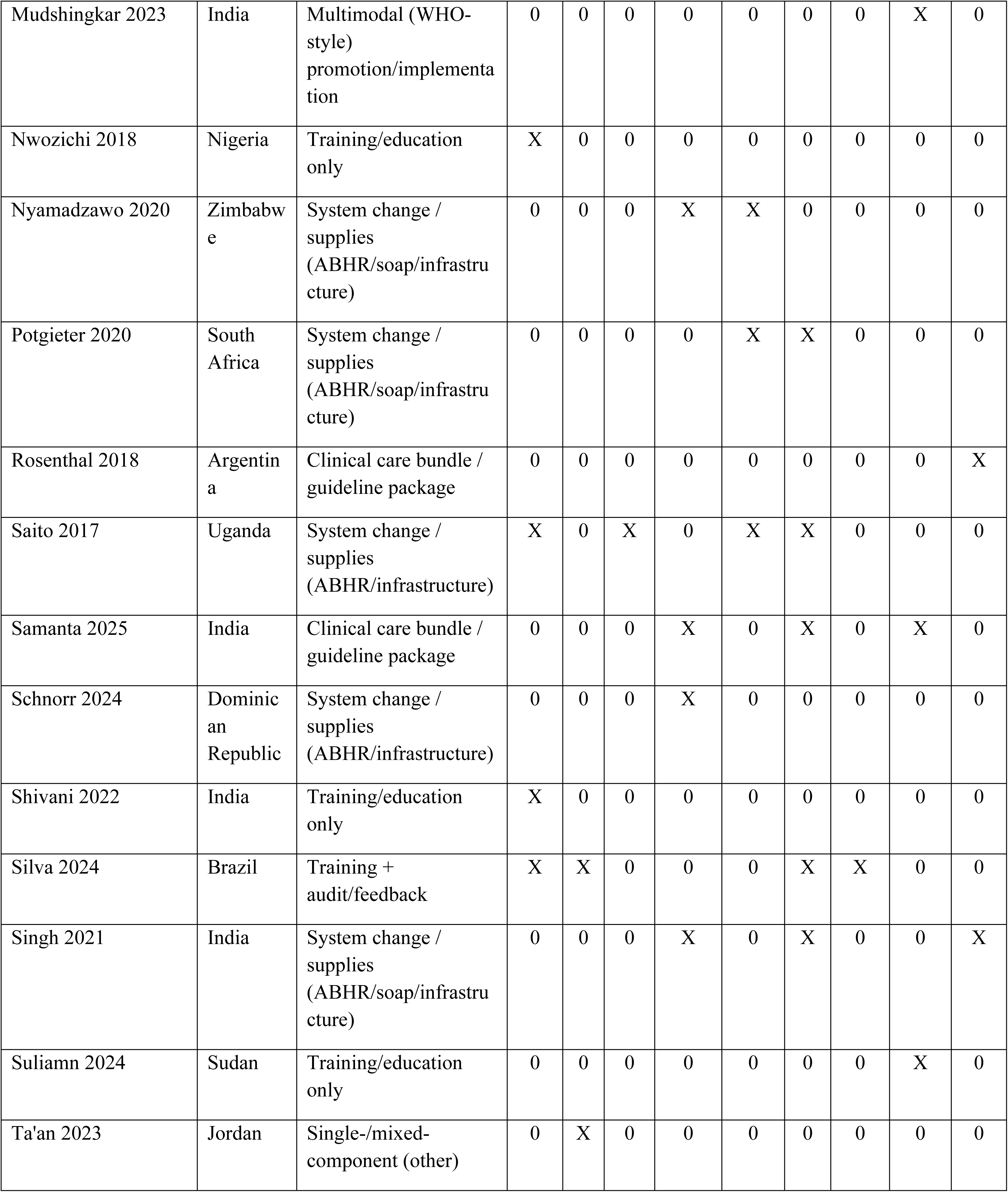
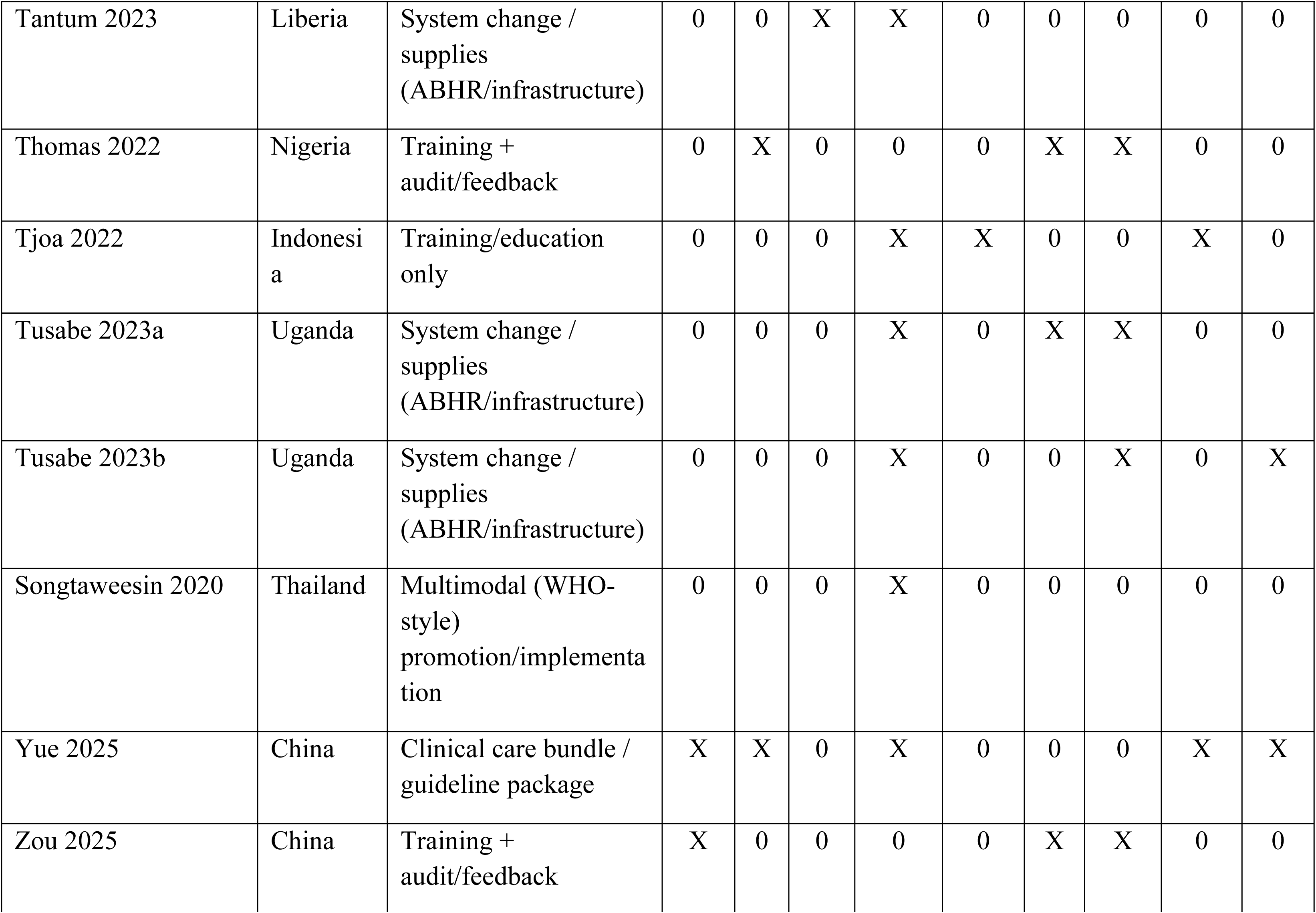
Barriers to implementing hand hygiene interventions across low- and middle-income countries (LMICs), by study, country, and intervention type. “X” indicates the barrier was reported; “0” indicates not reported/not mentioned.

**S Table 2.**
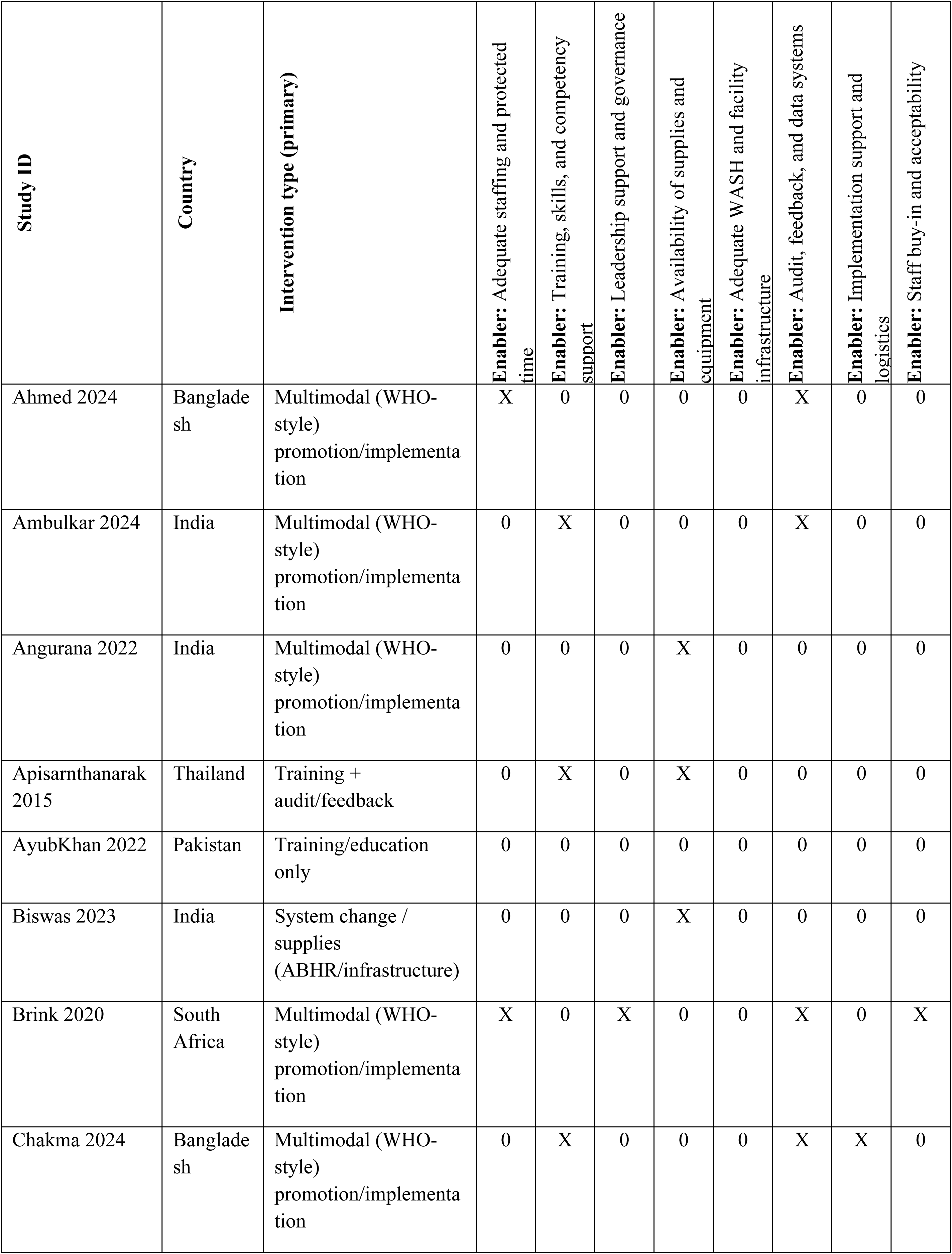
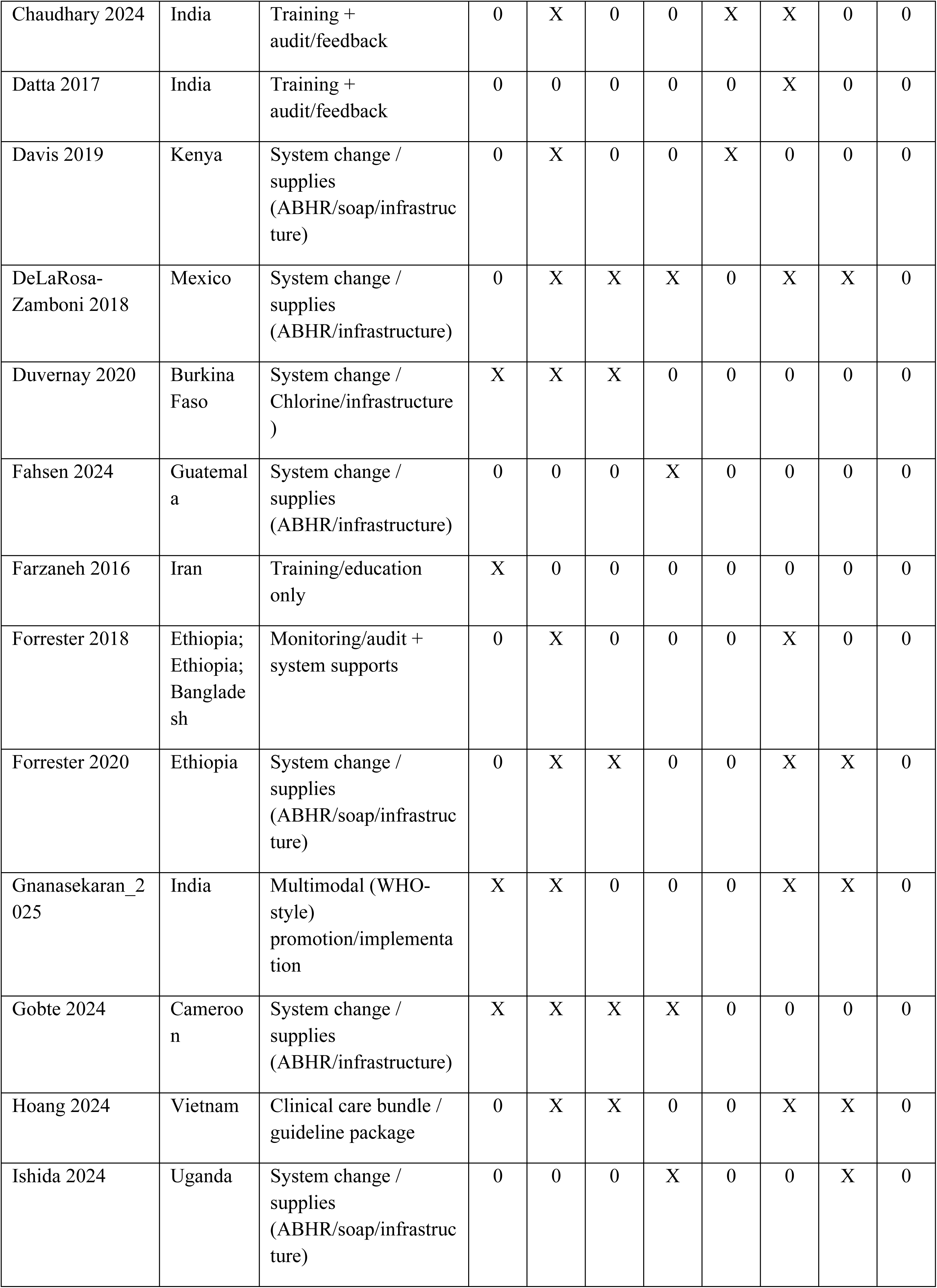
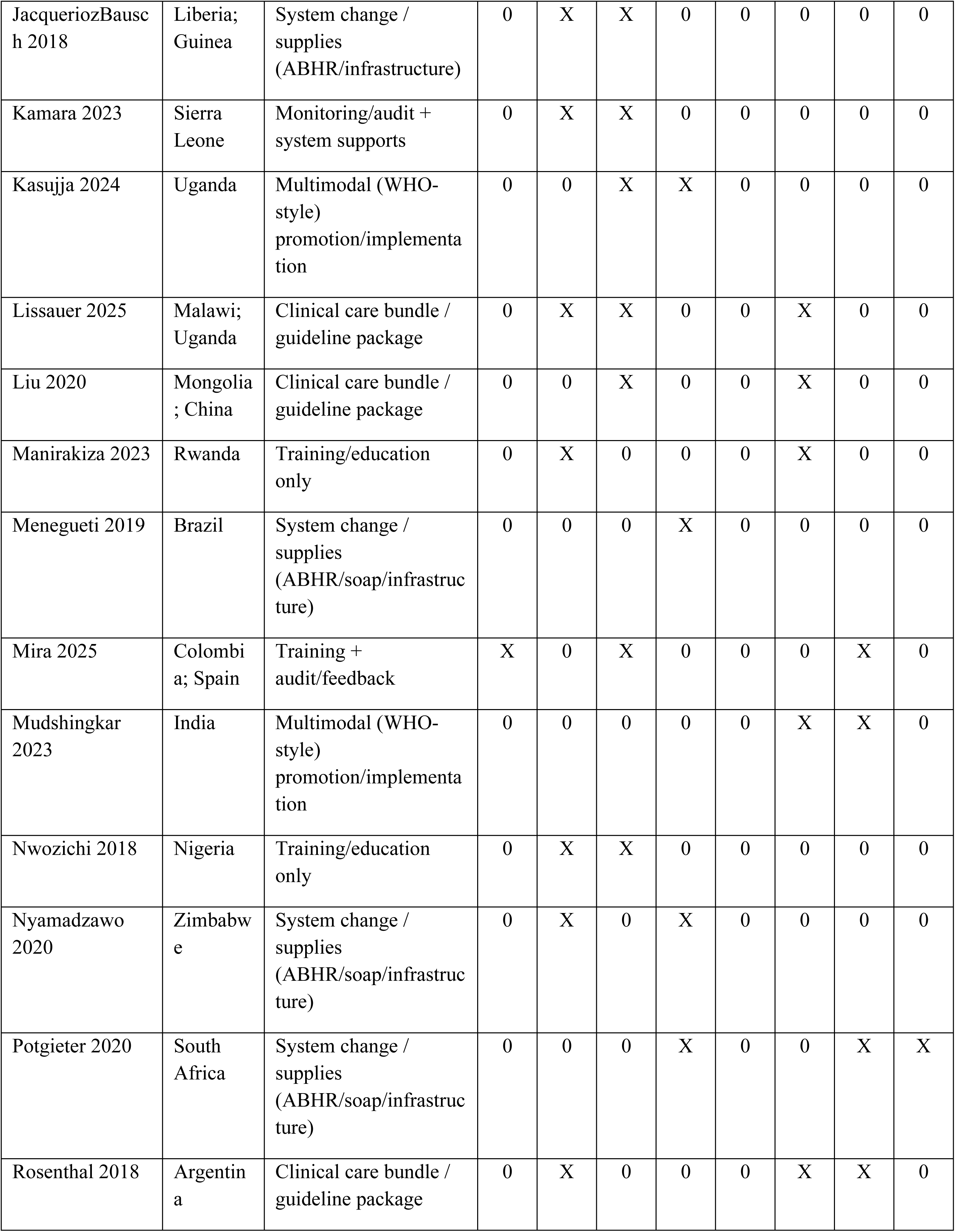
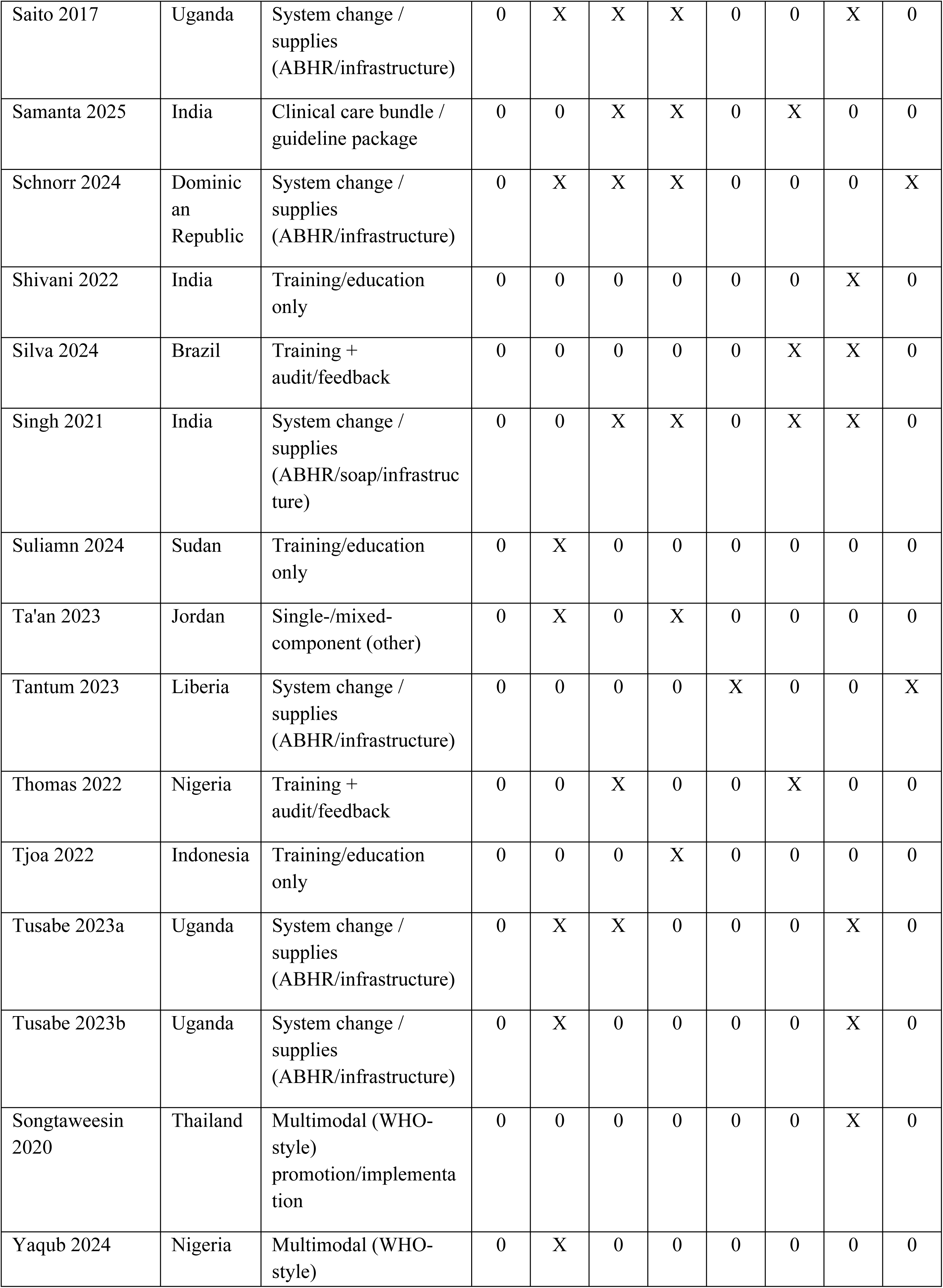
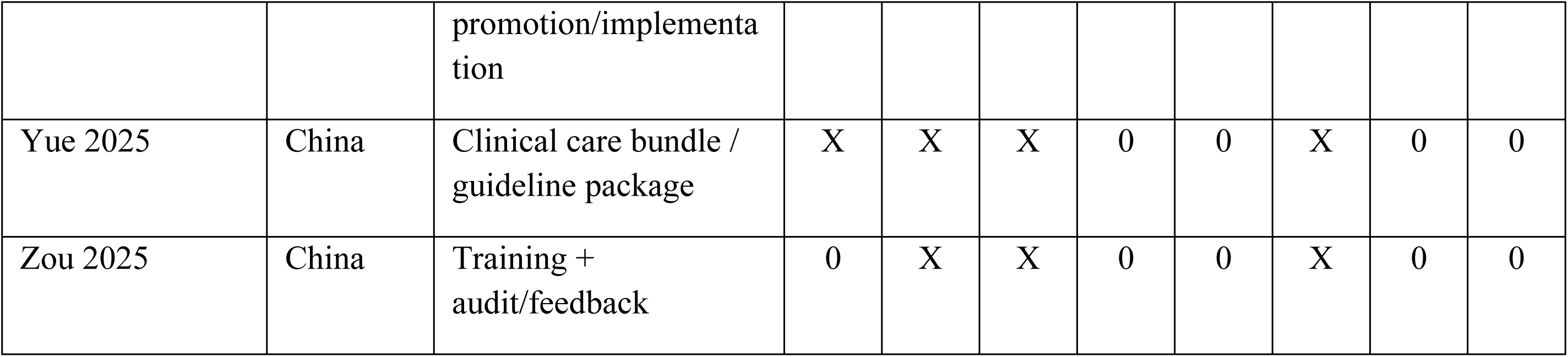
Enablers supporting implementation of hand hygiene interventions across low- and middle-income countries (LMICs), by study, country, and intervention type, mapped to predefined health system and behavioral domains. *Cells marked “X” indicate the enabler was reported; “0” indicates it was not reported or not mentioned*

**S Table 3.** Detailed characteristics of studies included in the systematic review.

| Study | Country | Study design | Intervention category | Intervention (summary) | Outcome(s) |
| --- | --- | --- | --- | --- | --- |
| Ahmed 2024[26] | Bangladesh | Pre–post intervention study. | Multimodal (WHO-style) promotion/implementation | Integrated IPC (infection prevention and control) + WASH (water, sanitation and hygiene) package implemented in all facilities: (1) IPC committee formation (facility manager as head; doctor as IPC focal point. | Hand hygiene resources/infrastructure |
| BenJmaa_2025[57] | Tunisia | Pre-post intervention study | Multimodal (WHO-style) promotion/implementation | Implementation of the WHO multimodal hand hygiene strategy. | Hand hygiene compliance and knowledge assessed via WHO questionnaire |
| McDavid_2025[22] | Belize | Pre–post intervention study | System change / supplies (ABHR/infrastructure) | Provision of ABHR wall mounts/bottles plus online hand hygiene training. | HH resources and hand hygiene adherence (HHA) |
| Ambulkar 2024[47] | India | Prospective, single-center interventional study | Multimodal (WHO-style) promotion/implementation | “Clean Cut” checklist-based, adaptive multimodal peri-operative IPC/QI package: surveillance + audits + directed at IPC training to improve compliance with 6 perioperative standards including hand hygiene. | Hand hygiene compliance; Healthcare-associated infections |
| Angurana 2022[58] | India | Quality improvement before-after study | Multimodal (WHO-style) promotion/implementation | Multimodal QI (PDSA): daily interactive teaching + fortnightly reinforcement; posters (5 moments + steps); confidential performance feedback using run charts; logistics/infrastructure: dedicated handwash corners w/ elbow taps, soap supply | Hand hygiene compliance |
| Apisarnthanarak 2015[59] | Thailand | Quasi-experimental pre–post | Training + audit/feedback | 3-arm behavior-based HH program (ICU-level allocation): S1 standard-of-care HH education every 3 months; S2 intensified HH interventions (monthly education emphasizing HH + patient safety and weekly workgroup discussions. | Hand hygiene compliance |
| AyubKhan 2022[60] | Pakistan | Mixed methods study | Training/education only | Interprofessional, task-based SSI-prevention training for staff (4 sessions over 2 months; every 2 weeks; 2 hours/session) using: (i) 10-min expert video (best practices), (ii) role plays (case scenarios covering antibiotic prophylaxis and hand hygiene). | Hand hygiene compliance; Knowledge/attitudes (KAP); Healthcare-associated infections |
| Biswas 2023[23] | India | Quasi-experimental Study | System change / supplies (ABHR/infrastructure) | Implemented site-specific elements: (1) System change: alcohol-based handrub (ABHR) dispensers at each bedside; (2) Training: 1-hour lecture delivered by consultant + infection control nurse (repeated across shifts). | Hand hygiene compliance; ABHR availability/use; Hand hygiene resources. |
| Brink 2020[46] | South Africa | Quasi-experimental study | Multimodal (WHO-style) promotion/implementation | Multi-modal, 5-phase organizational framework to improve hand hygiene (HH) ownership across a hospital group: executive governance + behavior campaign pledge; baseline situational analysis using WHO Hand Hygiene Self-Assessment Framework (HHSAF). | Hand hygiene compliance |
| Chakma 2024[50] | Bangladesh | Pre–post training intervention (no control group) | Multimodal (WHO-style) promotion/implementation | Multimodal HH training + reminders + supplies: 3-hour sessions over 5 days (lectures, video lessons, practical demonstrations incl. soap-and-water and alcohol-based hand rub), delivered by an IPC team; posters on WHO “Five Moments”; ensured HH supplies. | Hand hygiene compliance; Knowledge/attitudes (KAP) |
| Chaudhary 2024[37] | India | Prospective observation study | Training + audit/feedback | HH audits done by trained infection control nurses; audits included direct HH observation using WHO HH Observation forms "Awareness and reinforcement classes" for all cadres of workers; "continuous HH reinforcement activities in the hospital. | Hand hygiene compliance; HAIs |
| Datta 2017[61] | India | Quality improvement (QI) project using PDSA cycles | Training + audit/feedback | Hand hygiene training + monitoring using Glo Germ: Intensive handwashing sessions for mothers, healthcare providers, and ancillary workers. | Healthcare-associated infections |
| Davis 2019[29] | Kenya | Mixed methods study | System change / supplies (ABHR/soap/infrastructure) | WASH/hand-hygiene infrastructure package: installation of portable handwashing stations + drinking-water stations (metal stands; buckets with lids/taps; washbasins) plus starter soap and water treatment products. | Hand hygiene resources/infrastructure |
| DeLaRosa-Zamboni 2018[43] | Mexico | Quasi Experimental Study | System change / supplies (ABHR/infrastructure) | Monthly training for pediatric residents on HH and blood sampling, presentations on HCAI rates given to hospital leaders, HH posters in bathrooms but not clinical areas, ABHR on nursing trolleys and in every patientcare points. | Hand hygiene compliance; Knowledge/attitudes (KAP); HAIs; Product consumption/use rates |
| Duvernay 2020[62] | Burkina Faso | Non-random control pilot intervention | System change / Chlorine/infrastructure) | Electro-chlorinator (“WATA”) devices to produce sodium hypochlorite + operational training / hygiene sensitization. Four device types (WATA-Standard/Plus/Midi/Maxi) producing 5 g/L chlorine solution. | Healthcare-associated infections |
| Fahsen 2024[20] | Guatemala | Post Intervention Cross-sectional study design | System change / supplies (ABHR/infrastructure) | Project produced and distributed alcohol-based hand rub (ABHR) to 19 public primary/secondary HCFs in Quetzaltenango, Guatemala to increase HH access; repeated HH observations before (2021 baseline) and after (2022 follow-up) ABHR distribution. | Hand hygiene compliance; ABHR availability/use; Hand hygiene resources/infrastructure |
| Farzaneh 2016[51] | Iran | Quasi-experimental Study | Training/education only | Health Belief Model–based education for nurses to improve infection-control behaviors (handwashing, injections, dressings, standard precautions): four sessions (45 min each) delivered across work shifts; individual + group formats & face-to-face teaching. | Knowledge/attitudes (KAP) |
| Fofanah 2024[63] | Sierra Leone | Longitudinal hospital-based observation study | System change / supplies (ABHR/infrastructure) | Provision of locally produced ABHR following WHO formulation | ABHR availability/use |
| Forrester 2018[64] | Ethiopia;<br>Bangladesh | Prospective pre-/post-intervention observational QI study | Monitoring/audit + system supports | Clean Cut checklist-based surgical infection-prevention quality program: baseline observation + process mapping + feedback/brainstorming + locally led changes to improve adherence to perioperative IPC (incl. ABHR usage) | Hand hygiene related measures, ABHR availability/use; Hand hygiene resources/infrastructure; HAIs |
| Forrester 2020[65] | Ethiopia | Pre-post Prospective quality improvement study | Clinical care bundle / guideline package | Clean-Cut program targeting six standards: (1) hand and site hygiene (2) sterile field maintenance, (3) instrument sterilization, (4) antibiotic prophylaxis, (5) gauze counting Introduced alcohol-based hand rubs. | Hand hygiene compliance; HAIs |
| Gebremicael 2024[66] | Ethiopia | Pre-post intervention study | System change / supplies (ABHR/infrastructure) | Local production of ABHR, distribution of dispensers at every bed & staff education. | Hand hygiene compliance; ABHR availability/use |
| Gnanasekaran_2025[67] | India | pre-post interventional study | Multimodal (WHO-style) promotion/implementation | Supportive supervision + capacity building to improve WASH and IPC using the Water and Sanitation for Health Facility Improvement Tool (WASH FIT) and Kayakalp tool. | Hand hygiene compliance |
| Gobte 2024[25] | Cameroon | Pre-post intervention study | System change / supplies (ABHR/infrastructure) | Local production of ABHR. Wall mounted ABHR (produced in house) at point of care+ HH training, monitoring and feedback and HH champions. | Hand hygiene compliance; ABHR availability/use; HAIs |
| HajraMukasa 2023[68] | Uganda | Pre-post study design | Multimodal (WHO-style) promotion/implementation | At HCFs: Training and capacitating HCWs: On the job training and mentorship of HCWs on MNCH services; facilitating outreach by HCWs to provide MNCH services Construction of WASH facilities: Installing water systems to provide running water in maternity wards. | Hand hygiene resources/infrastructure |
| Hoang 2024[69] | Vietnam | Cross-sectional, longitudinal descriptive study | Clinical care bundle / guideline package | Ventilator Associated Pneumonia Care Bundle with 10 standards; 1) patient hand hygiene 2) elevated head 30-45 degrees, 3) oral hygiene (chlorhexidine), 4) stop sedation, 5) manage the breathing circuit. | Hand hygiene compliance |
| Ishida_2024[34] | Uganda | Post Intervention Cross-sectional study design | System change / supplies (ABHR/soap/infrastructure) | District-led production of WHO-formulation ABHR+ district-wide distribution to selected public/private not-for-profit HCFs; plus, one-time provision of portable handwashing stations to select HCFs. | Hand hygiene compliance; Hand hygiene resources/infrastructure; KAP; HAIs |
| JacquieriozBausch 2018[30] | Liberia; Guinea | Implementation case study | System change / supplies (ABHR/infrastructure) | Local capacity-building for ABHR production using WHO formula: IPC needs assessment by Ministry of Health validation/registration, selection of hospital-based production sites. | Hand hygiene compliance; Healthcare-associated infections |
| Kamara 2023[70] | Sierra Leone | "Before and after" observational study | Monitoring/audit + system supports | Following baseline HHSF, launched HH improvement initiative- "engagement of stakeholders at the national and hospital levels, actions to formalize patient engagement, and systematic audits of hand hygiene promotion resources" Ensure regular supply of ABHR. | Hand hygiene compliance; ABHR availability/use; Hand hygiene, HAIs |
| Kasujja 2024[24] | Uganda | Longitudinal before–after implementation | Multimodal (WHO-style) promotion/implementation | Implemented WHO multimodal hand hygiene (HH) improvement strategy using continuous quality improvement (CQI) (PDSA cycles), including governance/teams, system change (soap/ABHR availability), training/CME, monitoring + feedback & reminders/posters. | Hand hygiene compliance; Knowledge/attitudes (KAP); Healthcare-associated infections |
| Lindsjö 2015[71] | India | Pre-post intervention study | Multimodal (WHO-style) promotion/implementation | "WHO Hand Hygiene Intervention": distribution of ABHR (through placing dispensers in all wards), display of posters reminding HCWs to practice HH (on the wall near dispensers), continuing education for HCWs (monthly training sessions on appropriate use of ABHR). | ABHR availability/use; Healthcare-associated infections |
| Lissauer_2025[72] | Malawi; Uganda | Multicounty cluster-randomized trial | Clinical care bundle / guideline package | APT-Sepsis, a multicomponent facility-level implementation program including: “5 Moments for Hand Hygiene” with correct technique; (2) evidence-based maternal infection prevention/management and antibiotic use. | Hand hygiene compliance; ABHR availability/use; HAIs |
| Liu 2020[32] | Mongolia;<br>China | Before–after<br>observational study | Clinical care bundle /<br>guideline package | Ventilator care bundle (VCB), including head-of-bed elevation 30–45°, oral care with chlorhexidine 3–4x/day, adherence to aseptic technique + hand hygiene. | Hand hygiene compliance |
| Manirakiza 2023[73] | Rwanda | Prospective Cohort Study | Training/education only | Monthly neonatal nurse clinical mentorship (launched Aug 2021): 25 experienced neonatal nurses from referral hospitals oriented as mentors. | Knowledge/attitudes (KAP); Healthcare-associated infections |
| Meneguetti 2019[27] | Brazil | Cluster-randomized, double-blind, crossover study | System change / supplies (ABHR/soap/infrastructure) | Tested WHO EBHR (handrub) original formula with 1.45% glycerol and compared this to other concentrations (0, 0.5 and .75%) Participants used one formulation at a time for 7 days and then had an external observer evaluate their hands using WHO visual scale. | Hand hygiene compliance |
| Mira_2025[55] | Colombia | Quasi-experimental Study. | Training + audit/feedback | Brief virtual reality (VR) handwashing skills training (water + soap) aligned WHO recommendations: up to three 15-minute VR practice sessions with real-time visual/audio feedback + facilitator debrief. | Hand hygiene compliance; ABHR availability/use; (KAP); HAIs |
| Mudshingkar 2023[44] | India | pre–post Prospective interventional study. | Multimodal (WHO-style) promotion/implementation | Multicomponent HH improvement package in 6 ICUs: (1) Education/training (workshops, on-site demos, reminder posters); (2) Attitude/perception tools (display HH audit data in monthly HICC meetings; certify best-performing ICU; show pre/post hand imprints). | Hand hygiene compliance |
| Nwozichi 2018[74] | Nigeria | Pretest–posttest quasi-experimental | Training/education only | Structured infection-control training (4 weekly modules). Module 2 specifically covered hand hygiene, patient-care equipment, and preparation of JIK hypochlorite (3.5%) at different strengths; Module 3: PPE. | Knowledge/attitudes (KAP); Healthcare-associated infections |
| Nyamadzawo 2020[49] | Zimbabwe | Nonrandomized longitudinal (pre–post) interventional design | System change / supplies (ABHR/soap/infrastructure) | Pocket-sized 50 mL gel ABHR (Dettol; 69.4% denatured alcohol) provided to intervention nurses for point-of-care use + brief education on indications/amount. | Hand hygiene compliance; ABHR availability/use |
| Potgieter 2020[35] | South Africa | Non-randomized controlled before-and-after | System change / supplies (ABHR/soap/infrastructure) | Water-saving surgical hand preparation approaches: (1) Standard soap-and-water scrub under running tap; (2) Alcohol scrub method (brief prewash + alcohol-based scrub; alcohol between cases. | Hand hygiene resources/infrastructure |
| Rosenthal 2018[21] | Argentina | Prospective cohort before–after (pre–post surveillance) | Clinical care bundle / guideline package | INICC Multidimensional Approach (IMA) implemented via INICC Surveillance Online System (ISOS): (1) CLABSI prevention bundle, (2) education, (3) outcome surveillance, (4) process surveillance, (5) feedback on CLABSI rates/consequences, (6) performance feedback. | Hand hygiene compliance; Healthcare-associated infections |
| Saito 2017[75] | Uganda | Quasi-experimental study | System change / supplies (ABHR/infrastructure) | Locally made ABHR provision + hand hygiene (HH) education/training for healthcare providers (HCPs). Intervention phase included: (1) 1-L ABHR bottles mounted on ward walls in locally made holders + bottles on trolleys. | Hand hygiene compliance; ABHR availability/use; HAIs |
| Samanta 2025[31] | India | pre–post quality improvement evaluation study | Clinical care bundle / guideline package | Multidisciplinary quality improvement (QI) initiative using Plan–Do–Study–Act (PDSA) cycles to implement/refine a central line–associated bloodstream infection (CLABSI) prevention bundle, including Hand hygiene | Hand hygiene compliance; ABHR availability/use; HAIs |
| Schnorr 2024[19] | Dominican Republic | Longitudinal evaluation with repeated cross-sectional observation rounds | System change / supplies (ABHR/infrastructure) | Two-stage HH improvement package: (1) Local production + distribution of WHO-standard ABHR staff trained; ABHR bottled into 1-L pump dispensers mounted in patient-care areas; internal/external QC to ensure 70–80% alcohol. | Hand hygiene compliance; HAIs |
| Shivani 2022[76] | India | Quasi-experimental study | Training/education only | Reinforcement program for short peripheral catheter (SPC) initiation: (1) individual face-to-face interactive teaching including hand hygiene (40–45 min) + (2) group session on practical challenges/motivation; plus, IEC tools: adherence card attached to patient file. | Adherence to SPC care guideline steps |
| Silva 2024[38] | Brazil | Quasi-experimental Study | Training + audit/feedback | Implementation of the I-DECIDED assessment & decision tool for PIVC care in a pediatric inpatient unit. 20-min interactive education on the tool + discussion of barriers. | Hand hygiene compliance; Healthcare-associated infections |
| Singh 2021[40] | India | Prospective interventional study | System change / supplies (ABHR/soap/infrastructure) | QI package to increase correct point-of-care ABHR use alongside handwashing: staff education; poster reminder. | Hand hygiene compliance |
| Suliamn 2024[77] | Sudan | Pre-Post intervention study | Training/education only | Educational intervention; live presentation and video on proper glove donning; individual demonstrations of proper donning. | Knowledge/attitudes (KAP) |
| Ta'an 2023[78] | Jordan | Pre-post test experimental study. | Training/education only | Competency Outcomes and Performance Assessment (COPA)-based, competency-tailored infection prevention and control training for newly hired staff in a COVID-19 field hospital. Competency domains included hand and respiratory hygiene. | IPC Competency |
| Tantum 2023[54] | Liberia | Mixed-methods pilot study | System change / supplies (ABHR/infrastructure) | Personally worn hand hygiene holster (wearable device with 230 mL pump-style ABHR bottle) + hospital-based ABHR production to support refill. | Hand hygiene compliance; ABHR availability/use |
| Thomas 2022[28] | Nigeria | Pre-post-test study | Training + audit/feedback | Modular, self-paced, mobile-ready online course covering foundational infection prevention and control (IPC) concepts (10 modules; 10–20 min/module), using short vignette-based questions with feedback (formative assessment). Included a dedicated hand hygiene module. | Knowledge/attitudes (KAP) |
| Tjoa 2022[52] | Indonesia | Quasi-experimental, study | Training/education only | Online hand hygiene educational training (WHO-based module) The training was delivered twice (morning and afternoon) via zoom. | Hand hygiene compliance |
| Tusabe 2023a[41] | Uganda | Implementation evaluation study | System change / supplies (ABHR/infrastructure) | District-led, centralized local production + quality control + district-wide distribution of WHO ABHR to increase access at public healthcare facilities (and in Kasese also private) in two Western Uganda districts. | ABHR availability/Product consumption/use rates |
| Tusabe 2023b[42] | Uganda | Implementation case study | System change / supplies (ABHR/infrastructure) | District-wide establishment of local ABHR production units during health emergencies. | ABHR availability/use; Product consumption/use rates |
| Songtaweessin 2020[45] | Thailand | Pre-post intervention study | Multimodal (WHO-style) promotion/implementation | Multimodal hand hygiene promotion program including HH promotion videos sent to staff via mobile phone, HH signs at the bedside, distribution of portable alcohol gel bottles, HH promotion culture led by senior staff members and fob watch prizes. | Hand hygiene compliance; HAIs |
| Yaqub 2024[79] | Nigeria | Pre-Post intervention study | Multimodal (WHO-style) promotion/implementation | Modification to the environment; Visual reminders & provision of ABHR Modification of attitudes; HH BC workshop & verbal reminders Modification of technique; HH BC workshop & provision of ABHR | Hand hygiene compliance |
| Yue 2025[53] | China | Pre-post intervention study | Clinical care bundle / guideline package | Multi-year Plan-Do-Check-Act (PDCA) quality-improvement program to improve hand hygiene (HH) among healthcare workers (HCWs), informed by a staff survey and implemented as a bundled package: (1) regular HH training + assessments; (2) promotional campaigns. | Hand hygiene compliance; ABHR availability/use; Hand hygiene resources/infrastructure; (KAP); HAIs |
| Zou 2025[39] | China | Retrospective pre-vs post-intervention periods study | Training + audit/feedback | Cluster-based IPC interventions, including: (1) establishing a team of infection control observers (trained; supervision/management; reward & punishment system. | Hand hygiene compliance; HAIs |

**S Table 4.** Definitions of intervention categories used for coding included hand hygiene interventions, with illustrative examples of components assigned to each category.

| Intervention category | Definition used for coding | Examples of components |
| --- | --- | --- |
| **Multimodal (WHO-style) promotion/implementation** | Interventions combining multiple elements of the WHO Multimodal Hand Hygiene Improvement Strategy, delivered as an integrated package. These typically included three or more components addressing both behavior and system supports. | Combined training, point-of-care ABHR placement, reminders/posters, compliance monitoring, feedback, supervision, hand hygiene champions, leadership engagement. |
| **System change: ABHR availability, placement, or related infrastructure** | Interventions primarily focused on improving access to alcohol-based hand rub (ABHR) or related hand hygiene infrastructure at the point of care. | Installing ABHR dispensers, bedside ABHR placement, pocket bottles, local ABHR production, strengthening ABHR supply chains, dispenser maintenance, point-of-care access improvements. |
| **Training/education only** | Interventions primarily aimed at improving hand hygiene through knowledge or skills development, without major accompanying system-change or monitoring components. | Staff training sessions, workshops, demonstrations, refresher training, orientation sessions, educational materials, competency instruction. |
| **Training plus audit/feedback** | Interventions combining training or education with structured observation, monitoring, or performance feedback, but not meeting criteria for multimodal classification. | Training plus compliance audits, direct observation with feedback, performance review meetings, audit reports, feedback sessions. |
| **Clinical care bundle/guideline package** | Interventions in which hand hygiene was implemented as part of a broader clinical guideline, quality-improvement package, or care bundle rather than as a stand-alone hand hygiene strategy. | IPC bundles, device-care bundles, ward quality-improvement packages, guideline implementation packages, bundled workflow changes that included hand hygiene. |
| **System change: ABHR, soap, and hand hygiene infrastructure package** | Interventions focused on improving hand hygiene materials and infrastructure more broadly, including ABHR together with soap, water access, sinks, or related facilities. | ABHR plus soap provision, sink installation or repair, handwashing stations, water access improvements, dispenser systems, point-of-care hygiene materials package. |
| **Monitoring/audit plus system supports** | Interventions in which monitoring or audit was the dominant strategy, supported by enabling systems, but without the full range of multimodal components. | Observation tools, monitoring checklists, reporting systems, quality-improvement huddles, supervisory review, routine compliance tracking linked to support systems. |
| **System change: chlorine/infrastructure** | Interventions primarily focused on chlorine-based hand hygiene materials or related infrastructure, usually in outbreak or high-risk contexts. | Chlorine handwashing stations, chlorine solution provision, chlorine preparation systems, emergency or outbreak-response hand hygiene infrastructure. |

**S Table 5.** Predefined domains used to code implementation barriers and enablers for hand hygiene interventions.

| Predefined domain | Operational definition (what was coded) | Coding decision rule (how to assign) | Example barriers (what this looked like) | Example enablers (what this looked like) |
| --- | --- | --- | --- | --- |
| **Staffing, workload, and protected time** | Availability of sufficient staff and time to perform hand hygiene and to deliver intervention activities such as monitoring, feedback, supervision, and replenishment of supplies. | Code here when the determinant refers to staffing shortages, workload pressure, time constraints, turnover, competing clinical demands, or lack of protected time for IPC activities. | High workload, time pressure, staff shortages, rapid turnover, lack of time for observation or feedback, competing clinical priorities. | Protected time for IPC activities, adequate staffing, dedicated personnel, ward-level champions with time allocated, onboarding systems to manage turnover. |
| **Training, skills, and competency support** | Access to training, refresher education, coaching, mentorship, and competency support related to hand hygiene practice and intervention delivery. | Code here when the determinant relates to training quality, training frequency, refresher access, practical skill development, mentorship, or competency reinforcement. | Inadequate training, poor understanding of hand hygiene indications or technique, no refresher training, limited practical reinforcement. | Formal training packages, refresher sessions, bedside coaching, cascade training, mentorship, competency assessment and reinforcement. |
| **Leadership and governance** | Leadership engagement, governance structures, accountability, supervision, and management support for hand hygiene and IPC implementation. | Code here when the determinant describes leadership commitment, supervision, governance arrangements, accountability, managerial prioritization, or oversight of hand hygiene programs. | Weak leadership support, unclear accountability, inconsistent oversight, poor continuity under leadership change, limited governance for IPC. | Leadership sponsorship, visible management support, designated IPC focal persons, formal oversight structures, accountability mechanisms, champion systems. |
| **Supplies and equipment** | Availability, accessibility, placement, and maintenance of hand hygiene materials and related equipment, especially ABHR, soap, dispensers, and refill systems. | Code here when the determinant concerns stock-outs, supply reliability, point-of-care placement, equipment functionality, refill systems, or maintenance. | Stock-outs of ABHR or soap, broken dispensers, poor placement of supplies, interrupted supply chains, inadequate replenishment systems. | Reliable supply chains, point-of-care ABHR placement, dispensers or pocket bottles, regular replenishment, equipment maintenance, improved stock management. |

| **Predefined domain** | **Operational definition**<br>**(what was coded)** | **Coding decision rule**<br>**(how to assign)** | **Example barriers**<br>**(what this looked like)** | **Example enablers**<br>**(what this looked like)** |
| --- | --- | --- | --- | --- |
| **WASH and facility infrastructure** | Water, sanitation, sinks, handwashing stations, and broader physical infrastructure needed to support hand hygiene. | Code here when the determinant relates to water availability, sinks, handwashing stations, facility readiness, or physical infrastructure that supports or limits hand hygiene. | Intermittent water supply, nonfunctional sinks, inadequate handwashing stations, poor infrastructure at point of care, limited facility readiness. | Reliable water supply, functional sinks, handwashing stations near care areas, improved infrastructure, better point-of-care access conditions. |
| **Audit, feedback, and data systems** | Monitoring of hand hygiene practices and the use of audit data, feedback, reporting, and performance tracking to support implementation. | Code here when the determinant concerns routine monitoring, audit tools, observation systems, feedback loops, reporting, or data use for improvement. | Difficulty sustaining observations, incomplete documentation, irregular audits, lack of feedback to staff, data collected but not acted upon. | Structured observation systems, routine audit tools, feedback meetings, dashboards or reports, corrective action tracking, data used for improvement. |
| **Implementation support and workflow integration** | Practical arrangements that enable rollout and sustainment, including coordination, supervision, role clarity, workflow fit, and integration into routine care processes. | Code here when the determinant concerns operationalization of the intervention, implementation teams, supervision, role delineation, workflow integration, procurement coordination, or local support systems. | Poor coordination, unclear roles, weak supervision, intervention activities not integrated into routine workflow, logistical delays. | Implementation teams, supportive supervision, role clarity, workflow integration, coordinated distribution and replenishment, feasible operational routines. |
| **Staff buy-in and acceptability** | Perceived feasibility, acceptability, motivation, engagement, and norms among healthcare workers regarding hand hygiene practices or intervention components. | Code here when the determinant reflects motivation, resistance, staff engagement, perceived burden, acceptability of products or procedures, or social norms. | Low buy-in, resistance to intervention components, poor engagement with monitoring, perception that hand hygiene is low priority, reluctance to use ABHR consistently. | Staff engagement in planning, positive perceptions of ABHR or training, acceptance of monitoring and feedback, recognition or incentives, supportive norms. |
| **Financial support** | Funding availability and budgeting for recurrent consumables, training, monitoring systems, supervision, infrastructure maintenance, and program continuity. | Code here when the determinant is explicitly about budgets, financing, procurement funding, or ability to sustain inputs and activities. | No budget line for ABHR or soap, insufficient funds for monitoring or training, inability to maintain infrastructure or replace dispensers. | Dedicated budgets for supplies, financing for training and monitoring, planned replacement cycles, procurement support for continuity. |

**S Table 6.** Definitions and coding rules used for effect direction analysis.

| Effect direction category | Definition | Decision rules applied |
| --- | --- | --- |
| Improved | The outcome changed in a favorable direction following the intervention. | Classified as *improved* when: (i) a statistically significant favorable effect was reported (e.g., $p < 0.05$ or confidence interval excluding the null), **or** (ii) inferential statistics were not reported but absolute values (e.g., percentages, means, rates) showed a clear favorable change (e.g., increased hand hygiene compliance, improved knowledge scores, reduced HAI rates). |
| Worsened | The outcome changed in an unfavorable direction following the intervention. | Classified as *worsened* when: (i) a statistically significant unfavorable effect was reported, **or** (ii) inferential statistics were not reported but absolute values showed a clear unfavorable change (e.g., decreased compliance, increased HAI rates). |
| Mixed | The study reported inconsistent directions of effect within the same outcome domain. | Classified as *mixed* when a single study reported multiple relevant measures, subgroups, wards, or time points within an outcome domain that showed differing directions of effect (e.g., some indicators improved while others worsened or showed no change), preventing assignment of a single overall direction. |
| Unclear | Direction of effect could not be determined. | Classified as *unclear* when insufficient information was available to assign direction, including narrative statements without supporting numerical data, unclear reporting of pre/post or comparison group results, absence of statistical testing and absolute values, or when the direction of change was not explicitly stated. |

**S figure 1.**
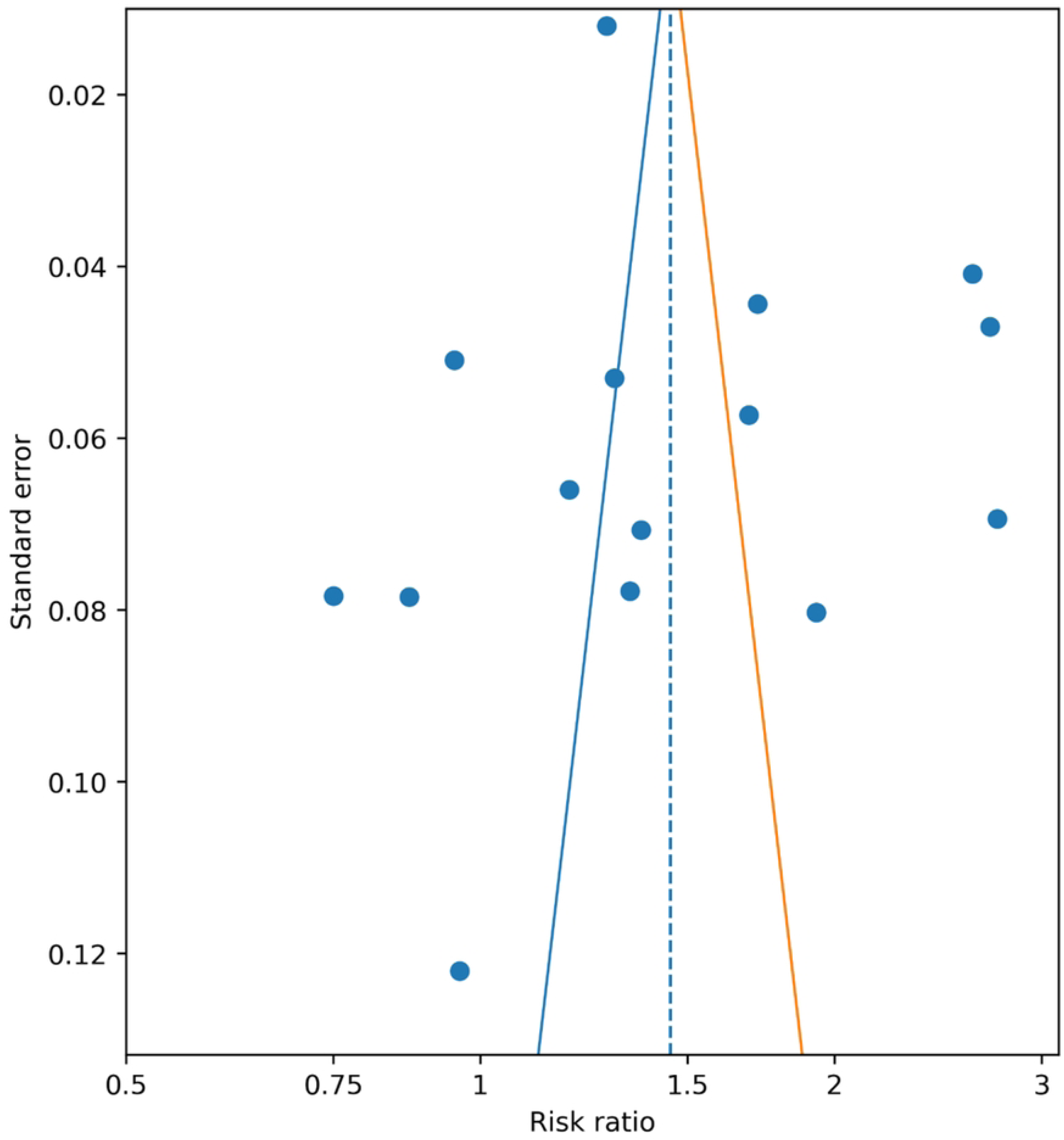
Funnel plot of studies included in the hand hygiene meta-analysis. Each point represents an individual study plotted according to its risk ratio and standard error. Funnel plot of studies included in the hand hygiene meta-analysis. Each point represents an individual study plotted according to its risk ratio and standard error. The dashed vertical line indicates the pooled risk ratio (RR = 1.45), and the diagonal lines represent the pseudo-95% confidence limits. Visual inspection suggests slight asymmetry; however, Egger’s test was not statistically significant (p = 0.437), indicating no strong evidence of small-study effects or publication bias.

**S figure 2.**
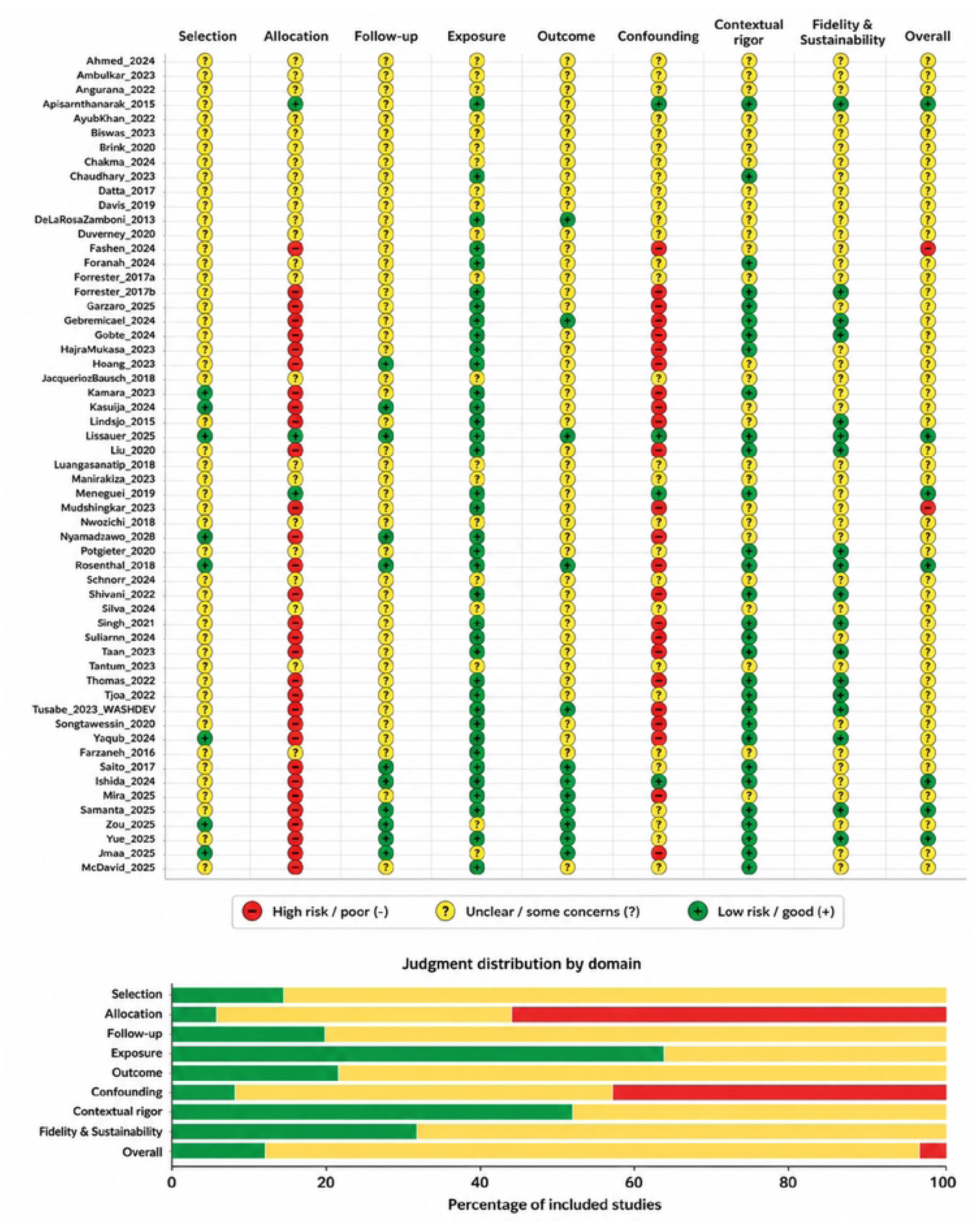
Methodological quality assessment of included studies. Traffic-light plot showing domain-level methodological quality judgments for each included study (top) and the distribution of judgments by domain (bottom). Green indicates low risk / good quality (+), yellow indicates unclear risk / some concerns (?), and red indicates high risk / poor quality (–). Domains assessed include selection, allocation, follow-up, exposure, outcome, confounding, contextual rigor, fidelity and sustainability, and overall methodological quality.

